# DXA-derived body composition and blood pressure in underweight and overweight adolescent girls in rural and urban South Africa

**DOI:** 10.64898/2026.09.15.26363182

**Authors:** SH Crouch, A Craig, DE Mathatha, LK Micklesfield, R Retnakaran, NJ Christofides, C Desmond, SJ Sharp, KK Ong, SM Tollman, K Kahn, SA Norris

## Abstract

**Background:** Elevated blood pressure (BP) during adolescence is an early marker of cardiovascular risk, particularly in settings experiencing under and overnutrition. However, the relationship between detailed body composition and BP across nutritional extremes remains poorly understood. We examined associations between dual-energy X-ray absorptiometry (DXA)-derived body composition and BP among underweight and overweight South African adolescent girls to assess multilevel correlates of BP.

**Methods:** The Ntshembo trial included 1268 girls 13-19-years from rural Agincourt and urban Soweto. Participants were selected as underweight or overweight. BP was measured in triplicate at a single visit and classified into clinic BP category ranges. DXA provided measures of fat mass (FM), fat-free soft tissue mass (FFSTM), and related indices. Results: Mean SBP and DBP were 110.0±11.2mmHg and 68.7±8.2mmHg, respectively; 21.1% of participants had elevated or hypertensive-range BP. Overweight adolescents had higher SBP and BP categories compared with underweight adolescents. DXA-derived adiposity measures were positively associated with SBP and BP category. FFSTM showed strong association with SBP. Higher FM/FFSTM was associated with elevated (RRR=4.25, 95%CI:1.77-10.20) and hypertensive-range BP (RRR=19.04, 95%CI:6.43–56.40). Caregiver hypertensive-range BP was independently associated with adolescent hypertensive-range BP (RRR=1.72-1.82). Sedentary behaviour independently associated with elevated BP, highlighting a modifiable behavioural target alongside intergenerational risk. Associations were more consistent for SBP than DBP.

**Conclusions:** both FM and FFSTM positively associated with BP, particularly SBP. Regional percentage analyses were consistent with higher BP when a greater proportion of total fat was distributed centrally rather than peripherally.

## Introduction

Adolescent elevated blood pressure (BP) is an increasingly important global public health concern, with growing recognition that cardiovascular risk has its origins early in the life course.^1^ BP in the elevated or hypertensive range during adolescence is an important early cardiovascular risk marker as BP in youth tracks into adulthood and is associated with later cardiovascular disease (CVD).^1, 2^ In low-and middle-income countries (LMICs) such as South Africa, this risk is amplified by rapid epidemiological transition, characterised by the coexistence of undernutrition, overnutrition, and rising cardiometabolic disease burden.^3^

While adiposity has long been implicated in the development of elevated BP in adolescence, evidence suggests that the relationship between body composition and BP is more nuanced than overall adiposity alone.^4^ Traditional anthropometric indicators such as body mass index (BMI) do not distinguish between fat mass (FM) and fat free soft tissue mass (FFSTM), a proxy for lean mass, nor do they capture regional fat distribution, which may differentially influence haemodynamic and metabolic pathways.^5^ Increasingly, research highlights that both FM (particularly central adiposity) and FFSTM are independently associated with BP, reflecting complex physiological pathways including vascular resistance, cardiac output, and metabolic demand.^6^ This complexity is especially relevant in South Africa with persisting undernutrition alongside rapidly increasing rates of overweight and obesity.^3^ This coexistence gives rise to distinct body composition phenotypes that may not be adequately captured by BMI alone, and which may confer differential cardiovascular risk.^7^ Underweight adolescents may have lower FFSTM and altered growth trajectories, whereas overweight adolescents may have excess adiposity and metabolic dysregulation; both phenotypes warrant investigation in settings experiencing the double burden of malnutrition, potentially providing useful information for targeted interventions. Furthermore, the distribution of adiposity (central vs peripheral) may be particularly important, given the established links with cardiometabolic dysfunction.^8^

Adolescent girls represent a population at heightened and distinct risk. Biological transitions during puberty, including hormonal changes and sex-specific fat deposition patterns, interact with behavioural and sociocultural factors to shape body composition and non-communicable disease risk profiles.^9–11^ In South Africa, these influences are further compounded by urban–rural disparities, differences in food environments, physical activity patterns, and socioeconomic conditions, all of which contribute to heterogeneity in body composition and BP outcomes.^9–11^

Despite this, there remains limited evidence examining how detailed body composition phenotypes relate to BP in adolescents, particularly across different BMI strata. The Ellisras Longitudinal Growth and Health Study showed that central adiposity, measured using skinfolds and waist circumference, was significantly associated with systolic BP (SBP) in rural adolescents.^12, 13^ However, the latter study did not explore differences by BMI status or incorporate precise measures of body composition. Importantly, both underweight and overweight adolescents may be vulnerable, albeit via distinct pathways, underweight individuals through reduced FFSTM and altered vascular development, and overweight individuals through excess adiposity and associated metabolic dysregulation ^12–14^ Addressing these gaps is critical for informing targeted interventions in populations experiencing the triple burden of malnutrition (underweight, overweight and micronutrient deficient).

Therefore, this study aims to investigate the association between dual-energy X-ray absorptiometry (DXA)-derived body composition phenotypes and BP among underweight and overweight South African adolescent girls in a rural and urban context. In addition, we assessed the contribution of demographic, household, behavioural, and caregiver-level factors to BP, allowing for a comprehensive understanding of multilevel correlates of adolescent BP. By integrating detailed body composition measures with contextual factors, this study provides important insight into early cardiovascular risk in a high-burden setting.

## Methods

### Study Population

Baseline data from the Ntshembo trial, a randomised controlled trial designed to assess the efficacy of a community health worker-administered intervention to address the burden of malnutrition (underweight and overweight) in adolescent girls, was analysed. Adolescent girls were classified as underweight or overweight according to International Obesity Task Force (IOTF) criteria and trial eligibility screening. Normal-weight adolescents were excluded; therefore, estimates should be interpreted as associations within nutritionally selected at risk groups rather than population prevalence estimates. Data were collected from 1268 adolescent girls aged 13-19 years. Trained research assistants who spoke the participant’s home language explained the study and all participants provided written informed consent or assent with caregiver consent if under 18 years, prior to participation. The Human Research Ethics Committee (Medical) at the University of the Witwatersrand approved the study (M211061).

### Study sites

Data were collected from two study sites, rural and urban, within South Africa. The rural study site was located in the Agincourt area of Bushbuckridge sub-district in northeast South Africa underpinned by a robust Health and socio-Demographic Surveillance System (HDSS) run by the SAMRC/Wits Rural Public Health and Health Transitions Research Unit since 1992. The study area, typical of marginalised former ‘homelands’, covers 450 km^2^ with some 117,000 people in 22,500 households distributed across 31 adjacent villages (260people/km^2^).^15^ The urban study site was located in Soweto, an urban-poor area of the city of Johannesburg covering 200 km^2^ with over 1.8 million people (6400people/km^2^). The SAMRC/Wits Ageing African Adult Research Unit has a research centre located the Chris Hani Baragwanath Academic hospital, at Africa’s largest hospital within Soweto and has 35 years of longitudinal research implementing cohorts, and experience of trials within the community.^16, 17^

### Demographic

A questionnaire was administered to primary caregivers to obtain household socio-demographics, education, employment and socio-economic data. A household asset score (an indicator of household socio-economic status; SES) was computed and used as an indicator of economic differentiation. The score comprised a count of thirteen household amenities and was subsequently categorised using a median split to distinguish between lower and higher asset households. A modified version of Household Food Insecurity Access Scale (HFIAS) was used to determine household food insecurity.^18^ Total HFIAS scores were used to categorise households as food secure (0–2), moderately at risk of food insecurity (3–5), food insecure (6– 10), or severely food insecure (11–27). Households were further categorised into a dichotomous variable of food secure or insecure for modelling purposes.

Adolescents completed a health survey to assess health behaviours and medical history, including self-reported tobacco use (smoking and vaping) classified as (never smoked, former smoker, or current smoker), previous hypertension diagnosis and current medication use. The Alcohol Use Disorders Identification Test (AUDIT) was used to determine risk of harmful alcohol consumption.^19^ The Pittsburgh Sleep Quality Index (PSQI) was used to assess self-report sleep quality. Sleep quality was scored on a scale of 0-21. Poor sleep quality was defined by a PSQI score of 5 or greater.^20^ The Patient Health Questionnaire-9 (PHQ-9) was used to assess the presence and severity of depression^21^. Probable depression was defined by a PHQ-9 score of 10 or greater or on medication. Dietary diversity was assessed using the Individual Dietary Diversity Questionnaire.^22^ Dietary practices were assessed using a modified food frequency questionnaire. For the analysis, consumption of each food group during the preceding 24 hours was coded as 1, while non-consumption was coded as 0. A dietary diversity score was calculated by summing the number of food groups consumed, yielding a maximum possible score of 14. Participants were then classified into tertiles according to their dietary diversity score: poor dietary diversity (≤5 food groups), average dietary diversity (6–7 food groups), and high dietary diversity (≥8 food groups). Sedentary behaviour was assessed using self-reported daily recreational screen time and categorised as <3 hours per day or ≥3 hours per day. Physical activity levels were determined using the Global School-based Student Health Survey Core Questionnaire Physical Activity Module,^23^ measuring the number of days per week on which participants engaged in at least 60 minutes of moderate-to-vigorous intensity physical activity (MVPA). Participants were categorised as inactive (0 days/week), insufficiently active (1–6 days/week), or active (7 days/week).

### Anthropometrics

Weight was measured using calibrated digital scales, and height was measured using a stadiometer with participants standing barefoot and maintaining an upright posture. Trained research assistants measured height and weight in triplicate to the nearest 0.1cm and 0.1kg using a portable stadiometer (SECA, Hamburg, Germany) and electronic scale (Omron body composition monitor), respectively in the adolescents and their caregivers. The average of the three measurements was used. BMI was calculated as weight (kg) / [height (m)]². For adolescents below the age of 18 years, sex and age-specific BMIs were determined and the International Obesity Task Force (IOTF) cut-offs were used to calculate these values accurately.^24^ For participants over 18 years of age BMI was classified as follows: underweight (<18.5 kg/m²), healthy weight (18.5-24.9 kg/m²), overweight (25.0-29.9 kg/m²), and obese (≥30 kg/m²).

### Blood Pressure

Brachial BP was measured using automated devices (HBP-1300, Omron Healthcare Co., LTD. Kyoto, Japan) in both adolescents and their caregivers. Following international protocol, participants were seated and asked to rest for at least 5-minutes prior to BP measurements. BP was measured in triplicate on the right arm in a quiet room with the monitor facing away from the participant, with a 2-minute rest interval between the measurements. The first BP measurement was discarded, and the average of the second and third measurements was used. If the second and third measures differ by >5mmHg, a fourth measure was performed, with the average of the two closest readings within a 5mmHg range used in the subsequent analyses. BP status of the adolescent participants, as all participants were aged 13–19 years, single-visit BP was classified using fixed thresholds aligned with the 2017 AAP adolescent scheme: normal, <120/<80 mmHg; elevated, 120–129 mmHg and <80 mmHg; stage 1 range, 130–139 mmHg or 80–89 mmHg; and stage 2 range, ≥140 mmHg or ≥90 mmHg. Stage 1 and stage 2 were combined as hypertensive-range BP for secondary analyses. These categories describe measurements from one visit and do not constitute a clinical diagnosis of hypertension..^25^ Adult (participants 18 years and above) BP status were classified into three BP categories: normotensive range BP (SBP <130 mmHg and DBP <85 mmHg), elevated range BP (SBP ≥130 mmHg and <140 mmHg and/or DBP ≥85 mmHg and <90 mmHg), or hypertensive range BP (SBP ≥140 mmHg and/or DBP ≥90 mmHg or self-reported hypertensive medication use).^26^ Mean Arterial Pressure (MAP) was calculated as MAP= (SBP + 2 x DBP)/3.

### Dual-energy x-ray absorptiometry (DXA)

Whole-body composition was assessed using dual-energy X-ray absorptiometry (DXA) at two geographically separate study sites. Two DXA scanners were used: a QDR 4500A DXA scanner (Hologic) with APEX System Software Version 4.6.0.2 in Soweto (urban), and a QDR DiscoveryA DXA scanner (Hologic) with APEX System Software Version 4.5.4 in Agincourt (rural). All scans were performed by trained research staff using standardised participant positioning and acquisition protocols across both sites to minimise inter-operator and inter-site variability. Routine quality control (QC) procedures were performed in accordance with the manufacturer’s recommendations before participant scanning. A phantom calibration scan was performed each morning before scanning commenced to verify instrument performance and calibration. In addition, a whole-body composition phantom scan was performed weekly. Participant scans were undertaken only after successful completion of the scheduled daily and weekly QC procedures. To assess comparability between the two DXA scanners, cross-calibration procedures were performed. Both scanners demonstrated acceptable short-term precision, with the magnitude of difference (0.23%) between the machines well below the ∼1.5% threshold applied for cross-calibration of Hologic instruments and within expected inter-scanner variation. As such, no cross-calibration adjustment was warranted. Whole body composition minus head (due to potential artifacts introduced by hair accessories) included whole body fat mass (WBFM), whole body lean mass (fat-free soft tissue mass), visceral adipose tissue (VAT), and subcutaneous adipose tissue (SAT). Regional fat distribution was expressed relative to WBFM with trunk fat (%FM) representing central fat distribution and limb fat (%FM) representing upper (arm) and lower (leg) body peripheral fat distribution, respectively. Fat mass index (FMI) and fat free soft tissue mass index (FFSTI, lean mass index) were calculated by dividing the respective mass values (in kg) by height squared (in m^2^). Prior to scanning, female participants of reproductive age were screened for pregnancy using a urine human chorionic gonadotrophin (hCG) test. Any participant with a positive pregnancy test was excluded from DXA scanning in accordance with radiation safety guidelines.

### Statistics

Study data were captured directly into and managed using REDCap (Research Electronic Data Capture) hosted at the University of Witwatersrand.^27, 28^ Data analysis was conducted using Stata v17. Data were assessed for normality using visual inspection of histograms, skewness and kurtosis, with normally distributed continuous variables summarised using means and standard deviations, and non-normally distributed variables using medians and interquartile ranges. Categorical data were presented as frequencies and percentages. The extent and pattern of missingness in all the variables used in the analysis was assessed. The proportion of missing data was low across variables, and missingness appeared to be random. A complete-case analysis approach was therefore adopted. The demographic and DXA body composition measures of the study population were compared across study site and BMI categories using chi-squared tests for categorical variables and analysis of variance (ANOVA) for continuous variables, with Bonferroni post hoc corrections for multiple comparison. To further explore differences between the four-study site BMI groups (rural underweight, rural overweight, urban underweight, and urban overweight), post-hoc pairwise chi-squared analyses were conducted. Associations between caregiver BP classification and adolescent BP classification were assessed.

Linear regression models were used first to estimate the associations of DXA body composition measures with SBP and DBP and then anthropometric and physiological characteristics and with SBP and DBP, after adjustment for age. Tanner stage was evaluated but not included in final models due to strong collinearity with age; age was retained as the primary adjustment variable to account for maturational effects. Multinomial logistic regression was used to determine the relative risk ratios between body composition measures and BP classification, adjusted for age, in the full sample of adolescents. Multinomial logistic regression was additionally used to examine associations between a range of demographic, household, behavioural, and caregiver-level factors and adolescent BP classification (normotensive [reference category], elevated, and hypertensive range BP). Results are presented as relative risk ratios (RRR) with corresponding 95% confidence intervals (CI) and p-values. A stepwise modelling approach was applied to explore the robustness of associations across adjusted models. Model 1 included demographic and household-level variables (age, study site, household socioeconomic status, number of people per sleeping room, and household food insecurity). Model 2 additionally incorporated adolescent behavioural factors, including BMI, tobacco smoking or vaping status, hazardous alcohol use, sedentary behaviour (screen time), physical activity level, sleep quality, dietary diversity, and depressive symptoms. Model 3 introduced caregiver-level factors, including caregiver age, caregiver BP classification, and caregiver BMI. Model 4 combined demographic, household, and adolescent behavioural variables to assess whether behavioural factors remained independently associated with BP classification after adjustment for contextual factors. Model 5 further added caregiver-level variables to the fully adjusted behavioural and household model. Model 6 extended Model 5 by replacing BMI with a direct measure of body composition (FMI derived from DXA), allowing assessment of whether associations with adiposity were robust to more precise body composition measurement. Lastly, Model 7 included the FM/FFSTM ratio derived from DXA, allowing simultaneous assessment of the roles of adiposity and lean mass.

To specifically assess the association between caregiver BP classification and the presence of elevated or hypertensive-range adolescent BP, adolescent BP classification was collapsed into a binary outcome (elevated or hypertensive-range BP vs normotensive BP). Modified Poisson regression with robust variance estimation was used to estimate prevalence ratios (PRs) and corresponding 95% confidence intervals (CI). This approach was selected because the outcome was common and prevalence ratios are more interpretable than odds ratios for cross-sectional data. Multicollinearity among independent variables was assessed using variance inflation factors (VIF). All variables demonstrated low collinearity, with a mean VIF of 1.11, indicating no evidence of problematic multicollinearity in the regression models. Analyses examining associations between DXA-derived body composition measures and BP outcomes (Tables 4 and 5) were conducted across multiple body composition indices and BP endpoints. These analyses were not corrected for multiple comparisons and should be interpreted as exploratory and hypothesis-generating rather than confirmatory. No a priori hypotheses were specified for individual body composition measures; rather, the analyses were designed to characterise the pattern of associations across the full range of body composition indices. Accordingly, p-values are reported without adjustment, and findings should be interpreted with appropriate caution. The consistency of associations across related measures and the magnitude of effect sizes were considered alongside statistical significance when drawing conclusions. Stratified analyses by BMI group and study site are presented for description only. As stratified analyses cannot establish effect modification (a significant association in one stratum and a non-significant association in another does not constitute a formal interaction), effect modification by BMI group and by study site was tested directly using interaction terms (body-composition index × moderator) in the full sample, adjusted for adolescent age and the other moderator. Interactions with blood-pressure classification were assessed by likelihood-ratio test. Given the number of interaction tests across body-composition indices, outcomes, and two moderators, we applied a Benjamini–Hochberg false discovery rate (FDR) correction across the whole family of interaction tests. Stratified estimates are therefore presented for description only. Of 60 interaction tests, 16 were nominally significant (p<0.05); the nominal signals were concentrated in interactions between adiposity measures (subcutaneous and visceral adipose tissue, fat mass, fat mass index, and regional fat) and study site, with associations tending to be stronger at the rural site. However, no interaction remained significant after Benjamini-Hochberg correction (all q>0.05; smallest q≈0.07). Differences in the strength of body composition and blood-pressure associations across BMI groups or study sites should therefore be regarded as exploratory.

## Results

The study population comprised 1268 adolescent girls (mean age 15.5±1.5 years), of whom 671 were from the rural study site (Agincourt) and 597 from the urban study site (Soweto). Urban adolescents were significantly older than their rural counterparts (mean age: urban underweight 15.8±1.7 years, urban overweight 16.1±1.6 years vs. rural underweight 14.9±1.3 years, rural overweight 15.2±1.3 years). Across the cohort, 43.5% were classified as underweight and 56.5% as overweight. Anthropometric and physiological characteristics are presented in **Table 1**. Mean SBP in the cohort was 110.0±11.2 mmHg and mean DBP was 68.7±8.2 mmHg, and 8% of the whole sample were classified as hypertensive. Both SBP and DBP were higher among overweight adolescents compared to their underweight counterparts, in both study sites. There was a higher prevalence of both elevated and hypertensive range BP in the overweight adolescents compared to the underweight adolescents. DXA-derived measures of body composition showed clear differences by BMI status and study site. Overweight adolescents had higher total FM, FMI, and central adiposity measures, including VAT, SAT, compared to underweight adolescents. Urban overweight participants had the highest levels of central adiposity across all measures, while FFSTI was also higher among overweight adolescents but showed less variation by study site. Behavioural characteristics also differed by study site (Table 1). Urban adolescents were more physically active, with a lower proportion classified as inactive (62.8-66.8%) compared to rural adolescents (78.1-83.0%). Conversely, urban adolescents had higher dietary diversity, with over 42% reporting high dietary diversity compared to fewer than 9% of rural adolescents. The prevalence of tobacco use, harmful alcohol consumption, depressive symptoms, and poor sleep quality was also higher among urban participants. Household socioeconomic and caregiver characteristics differed by study site and BMI status (**Table 2**). Household SES was generally higher among urban participants, while food insecurity was prevalent overall (62.2%), but higher in urban groups. Caregiver employment patterns also differed, with rural caregivers more likely to have never been employed compared to urban caregivers.

**Table 1.** Anthropometry, physiological and behavioural characteristics of adolescent girls.

| Variable | Total<br>(n=1268) | Rural<br>(n=671) |  | Urban<br>(n=597) |  |
| --- | --- | --- | --- | --- | --- |
|  |  | Underweight<br>(n=347) | Overweight<br>(n=324) | Underweight<br>(n=205) | Overweight<br>(n=392) |
| Adolescent age (years) | 15.5 (1.5) | 14.9 (1.3)<sup>a,b</sup> | 15.2 (1.3)<sup>c,d</sup> | 15.8 (1.7)<sup>a,c</sup> | 16.1 (1.6)<sup>b,d</sup> |
| Weight (kg) | 54.8 (13.1) | 41.3 (4.27)<sup>a,b</sup> | 65.7 (7.0)<sup>a,c</sup> | 41.8 (4.5)<sup>c,d</sup> | 64.7 (6.4)<sup>b,d</sup> |
| Height (cm) | 158.3 (6.4) | 159.0 (6.6)<sup>a</sup> | 159.5 (6.2)<sup>b,c</sup> | 158.0 (6.9)<sup>b</sup> | 156.8 (5.9)<sup>a,c</sup> |
| BMI (kg/m<sup>2</sup>) | 21.7 (5.0) | 16.3 (0.87)<sup>a</sup> | 25.4 (1.7)<sup>a</sup> | 16.7 (1.0)<sup>a</sup> | 26.3 (1.6)<sup>a</sup> |
| SBP (mmHg) | 110.0 (11.2) | 106.0 (10.4)<sup>a</sup> | 115.5 (10.5)<sup>a,b</sup> | 104.5 (10.4)<sup>b,c</sup> | 111.8 (10.5)<sup>a,c</sup> |
| DBP (mmHg) | 68.7 (8.2) | 66.5 (7.6)<sup>a</sup> | 70.2 (7.8)<sup>a,b</sup> | 67.7 (8.1)<sup>b,c</sup> | 69.5 (8.7)<sup>a,c</sup> |
| Mean Arterial Pressure | 82.4 (8.4) | 79.7 (7.6)<sup>a,b</sup> | 85.3 (7.6)<sup>a</sup> | 79.9 (8.3) | 83.8 (8.5)<sup>b</sup> |
| Blood Pressure Classification |  |  |  |  |  |
| Normotensive | 1000 (78.9) | 306 (88.2)<sup>a</sup> | 214 (66.0)<sup>a,b</sup> | 179 (87.3)<sup>b,c</sup> | 301 (76.8)<sup>a,b,c</sup> |
| Elevated BP | 166 (13.1) | 30 (8.6) | 73 (22.5) | 17 (8.3) | 46 (11.7) |
| Hypertensive | 102 (8.0) | 11 (3.2) | 37 (11.5) | 9 (4.4) | 45 (11.5) |
| VAT area (cm<sup>2</sup>) | 36.6 (22.0) | 17.49 (6.6)<sup>a</sup> | 38.25 (13.4)<sup>a</sup> | 21.82 (7.9)<sup>a</sup> | 60.47 (18.7)<sup>a</sup> |
| SAT area (cm<sup>2</sup>) | 227.2 (103.4) | 127.4 (22.0)<sup>a</sup> | 276.1 (66.9)<sup>a</sup> | 134.0 (22.5)<sup>a</sup> | 326.3 (71.4)<sup>a</sup> |
| VAT:SAT | 0.16 (0.05) | 0.14 (0.04)<sup>a</sup> | 0.14 (0.04)<sup>b</sup> | 0.16 (0.04)<sup>a,b</sup> | 0.19 (0.05)<sup>a,b</sup> |
| Whole body fat mass (kg) | 18.3 (7.8) | 10.3 (2.2)<sup>a</sup> | 23.0 (4.6)<sup>a</sup> | 11.1 (2.3)<sup>a</sup> | 25.5 (4.3)<sup>a</sup> |
| Whole body fat mass (% WBM) | 32.1 (7.5) | 25.0 (4.6)<sup>a</sup> | 34.8 (4.7)<sup>a</sup> | 26.5 (4.1)<sup>a</sup> | 39.3 (4.1)<sup>a</sup> |
| Whole body FFSTM (kg) | 30.4 (5.3) | 27.8 (3.4)<sup>a</sup> | 35.9 (4.2)<sup>a</sup> | 24.4 (3.0)<sup>a</sup> | 31.2 (3.3)<sup>a</sup> |
| Fat Mass Index (FMI, kg/m<sup>2</sup>) | 7.3 (3.1) | 4.1 (0.8)<sup>a</sup> | 9.0 (1.6)<sup>a</sup> | 4.5 (0.8)<sup>a</sup> | 10.4 (1.5)<sup>a</sup> |
| Fat Free Soft Tissue Mass Index (FFSTI, kg/m<sup>2</sup>) | 12.1 (1.8) | 11.0 (0.9)<sup>a</sup> | 14.1 (1.1)<sup>a</sup> | 9.7 (0.7)<sup>a</sup> | 12.7 (0.9)<sup>a</sup> |
| Trunk fat (% WBFM) | 39.4 (3.8) | 37.4 (2.8)<sup>a</sup> | 39.6 (3.7)<sup>a</sup> | 38.5 (3.4)<sup>a</sup> | 41.6 (3.6)<sup>a</sup> |
| Limb fat (% WBFM) | 60.6 (3.8) | 62.6 (2.8)<sup>a</sup> | 60.4 (3.7)<sup>a</sup> | 61.5 (3.4)<sup>a</sup> | 58.4 (3.6)<sup>a</sup> |
| Arm fat (% WBFM) | 11.2 (1.4) | 10.8 (1.1)<sup>a,b</sup> | 12.3 (1.1)<sup>a,c</sup> | 9.7 (1.2)<sup>c</sup> | 11.6 (1.1)<sup>b,c</sup> |
| Leg fat (% WBFM) | 49.3 (4.3) | 51.8 (3.0)<sup>a</sup> | 48.1 (4.0)<sup>a</sup> | 51.8 (3.5)<sup>a</sup> | 46.8 (3.9)<sup>a</sup> |
| Whole body fat mass (kg)/ FFSTM (kg) ratio | 0.59 (0.21) | 0.37 (0.07)<sup>a</sup> | 0.64 (0.11)<sup>a</sup> | 0.46 (0.10)<sup>a</sup> | 0.82 (0.13)<sup>a</sup> |
| Depression (freq. (yes; %)) | 357 (28.2) | 73 (21.0)<sup>a,b</sup> | 71 (21.9)<sup>c,d</sup> | 80 (39.0)<sup>a,c</sup> | 133 (33.9)<sup>b,d</sup> |
| Tobacco or vape smoker (yes; freq. (%)) |  |  |  |  |  |
| Never | 1163(91.7) | 347 (100.0)<sup>a,b</sup> | 323 (99.7)<sup>c,d</sup> | 165 (80.5)<sup>a,c</sup> | 328 (83.7)<sup>b,d</sup> |
| Former smoker | 55 (4.3) | 0 (0.0) | 1 (0.3) | 16 (7.8) | 38 (9.7) |
| Current smoker | 50 (3.9) | 0 (0.0) | 0 (0.0) | 24 (11.7) | 26 (6.6) |
| Harmful alcohol consumption |  |  |  |  |  |
| No risk | 1242 (98.0) | 347 (100.0)<sup>a,b</sup> | 321 (99.1)<sup>c,d</sup> | 198 (96.6)<sup>a,c</sup> | 376 (95.9)<sup>b,d</sup> |
| Risk | 26 (2.0) | 0 (0.0) | 3 (0.9) | 7 (3.4) | 16 (4.1) |
| Sedentary behaviour (freq. (%)) |  |  |  |  |  |
| <3 hours per day | 444 (35.0) | 125 (36.0) | 108 (33.3) | 79 (38.5) | 132 (33.7) |
| ≥3 per day | 824 (65.0) | 222 (64.0) | 216 (66.7) | 126 (61.5) | 260 (66.3) |
| Physical activity (freq. (%)) |  |  |  |  |  |
| Inactive | 924 (72.9) | 288 (83.0)<sup>a,b</sup> | 253 (78.1)<sup>c,d</sup> | 137 (66.8)<sup>a,c</sup> | 246 (62.8)<sup>b,d</sup> |
| Insufficiently Active | 293 (23.1) | 42 (12.1) | 51 (15.7) | 66 (32.2) | 134 (34.2) |
| Active | 51 (4.0) | 17 (4.9) | 20 (6.2) | 2 (1.0) | 12 (3.1) |
| Sleep quality (freq. (%)) Poor | 82 (6.5) | 6 (1.7) | 8 (2.5) | 23 (11.2) | 45 (11.5) |
| Dietary diversity (freq. (%)) |  |  |  |  |  |
| Poor (≤5 food groups) | 700 (55.5) | 227 (65.4)<sup>a,b</sup> | 212 (65.4)<sup>c,d</sup> | 88 (42.9)<sup>a,c</sup> | 173 (44.1)<sup>b,d</sup> |
| Average (6-7) | 256 (20.3) | 92 (26.5) | 81 (25.0) | 30 (14.6) | 53 (13.5) |
| High (≥8) | 306 (24.1) | 25 (7.2) | 28 (8.6) | 87 (42.4) | 166 (42.3) |

Items with the same superscript letters denote proportions differ significantly from each other at the $p < 0.05$ level.

**Table 2.** Household socio-economic and caregiver demographic characteristics.

| Variable | Total<br>(n=1268) | Rural<br>(n=671) |  | Urban<br>(n=597) |  |
| --- | --- | --- | --- | --- | --- |
|  |  | Underweight<br>(n=347) | Overweight<br>(n=324) | Underweight<br>(n=205) | Overweight<br>(n=392) |
| Household SES index | 8.7 (2.7) | 8.2 (2.2)<sup>a,b</sup> | 8.9 (3.1)<sup>c</sup> | 8.9 (3.1)<sup>a</sup> | 9.2 (3.1)<sup>b,c</sup> |
| Food Security |  |  |  |  |  |
| Food Secure | 469 (37.0) | 152 (43.8)<sup>a,b</sup> | 151 (46.6) | 55 (26.8)<sup>a</sup> | 111 (28.3)<sup>b</sup> |
| At Risk/Food Insecure | 771 (60.8) | 180 (51.9) | 163 (50.3) | 149 (72.7) | 179 (71.2) |
| Declined to answer | 28 (2.2) | 15 (4.3) | 10 (3.1) | 1 (0.5) | 2 (0.5) |
| Caregiver Age | 40.0 (34.0; 49.0) | 40.00 (33.0; 50.0) | 41.00 (34.0; 51.0) | 40.0 (33.0; 47.0) | 41.0 (35.0; 47.0) |
| Caregiver Education |  |  |  |  |  |
| Some Schools | 775 (60.7) | 195 (58.4)<sup>a</sup> | 175 (55.7)<sup>b</sup> | 136 (66.3)<sup>a,b</sup> | 249 (63.9) |
| Completed Secondary Schooling | 407 (32.7) | 123 (36.8) | 115 (36.6) | 51 (24.9) | 118 (30.3) |
| Completed Tertiary Schooling | 81 (6.5) | 16 (4.8) | 24 (7.6) | 18 (8.8) | 23 (5.9) |
| Caregiver Employment |  |  |  |  |  |
| Never Employed | 631 (49.8) | 263 (75.8)<sup>a,b</sup> | 238 (73.5)<sup>c,d</sup> | 48 (23.4)<sup>a,c</sup> | 82 (20.9)<sup>b,d</sup> |
| Not Currently Employed | 346 (27.3) | 29 (8.4) | 29 (9.0) | 107 (52.5) | 181 (46.2) |
| Currently Employed | 269 (21.2) | 48 (14.8) | 48 (14.8) | 50 (24.4) | 128 (32.7) |
| Declined to answer | 22 (1.7) | 9 (2.8) | 9 (2.8) | 0 (0.0) | 1 (0.3) |
| Caregiver SBP (mmHg) | 128.8 (17.8) | 127.4 (11.6) | 128.46 (11.9) | 129.76 (23.8) | 129.71 (21.9) |
| Caregiver DBP (mmHg) | 83.7 (11.7) | 82.9 (9.5) | 84.2 (9.1) | 83.47 (13.7) | 84.05 (14.0) |
| Caregiver Mean Arterial Pressure | 98.7 (13.0) | 97.7 (9.2) | 98.94 (9.1) | 98.90 (16.7) | 99.27 (15.9) |
| Caregiver Blood Pressure |  |  |  |  |  |
| Normotensive | 617 (48.7) | 178 (51.3) | 146 (45.1)<sup>a</sup> | 107 (52.2)<sup>a</sup> | 186 (47.4) |
| Elevated BP | 233 (18.4) | 66 (19.0) | 71 (21.9) | 27 (13.2) | 69 (17.6) |
| Hypertensive | 389 (30.7) | 89 (26.6) | 96 (29.6) | 69 (33.7) | 135 (34.4) |
| Missing | 29 (2.3) | 14 (4.0) | 11 (3.4) | 2 (1.0) | 2 (0.5) |
| Caregiver BMI (kg/m<sup>2</sup>) | 29.6 (6.5) | 27.83 (4.2)<sup>a,b</sup> | 30.2 (4.9)<sup>a,c</sup> | 26.8 (7.4)<sup>c,d</sup> | 31.20 (8.0)<sup>b,d</sup> |
| BMI Classification |  |  |  |  |  |
| Caregiver underweight | 19 (1.50) | 3 (0.86)<sup>a</sup> | 0 (0.0)<sup>a</sup> | 9 (4.4)<sup>a</sup> | 7 (1.8)<sup>a</sup> |
| Caregiver healthy weight | 273 (21.53) | 82 (23.6) | 35 (10.8) | 63 (30.7) | 93 (23.7) |
| Caregiver overweight | 453 (35.73) | 159 (45.8) | 143 (44.1) | 62 (30.2) | 89 (22.7) |
| Caregiver obese | 523 (41.25) | 103 (29.7) | 146 (45.0) | 71 (34.6) | 203 (51.8) |

Items with the same superscript letters denote proportions differ significantly from each other at the 0.05 level.

BMI classification showed significant differences across groups (**Table 2**). Among rural overweight adolescents, 45.0% had caregivers classified as obese, compared to 29.7% among rural underweight adolescents. In the urban site, 51.8% of overweight adolescents had obese caregivers compared to 34.6% of underweight adolescents. Conversely, underweight adolescents were more likely to have caregivers within the healthy weight range (rural: 23.6%; urban: 30.7%). Caregiver BP classification did not differ significantly between underweight and overweight adolescent groups within either study site. The prevalence of caregiver hypertension ranged from 26.6% (rural underweight) to 34.4% (urban overweight), with no statistically significant pairwise differences.

We assessed the concordance of BP classification between adolescents and their caregivers (**Table 3**). Overall, 46.4% of caregiver-adolescent dyads were classified in the same BP category. In contrast, 11.2% of dyads showed a higher BP category in the adolescent compared with the caregiver, and 42.4% of adolescents were in a lower BP category than their caregivers. Among adolescents with hypertensive caregivers, 21.9% had either elevated or hypertensive range BP. Finally, among hypertensive range BP adolescents, 43.9% had caregivers who were also in the hypertensive range BP. When stratified by nutritional status and study site, distinct patterns emerged. Among underweight adolescents, both rural and urban, the overall prevalence of elevated or hypertensive-range BP was low regardless of caregiver BP status. In the rural underweight group, the proportion of adolescents with elevated or hypertensive-range BP was similar across caregiver BP categories: 12.2% among those with normotensive caregivers, 12.2% with elevated BP caregivers, and 11.3% with hypertensive caregivers. Similarly, in the urban underweight group, the prevalence of elevated or hypertensive-range BP remained low (12.2%,7.4% and 16% among those with normotensive, elevated BP and hypertensive caregivers respectively), though 88.9% of the small number of urban underweight adolescents classified as hypertensive had caregivers who were also hypertensive. In the rural overweight group, 38.3% of adolescents with normotensive caregivers had elevated or hypertensive-range BP, while 27.1% of those with hypertensive caregivers had elevated or hypertensive-range BP. In the urban overweight group, 28.1% of adolescents with hypertensive caregivers had elevated or hypertensive-range BP, compared to 18.0% of those with normotensive caregivers. Among urban overweight adolescents classified as hypertensive, 44.4% had hypertensive caregivers. To formally quantify the strength of these intergenerational associations, we used modified Poisson regression to estimate the prevalence of elevated or hypertensive-range BP in adolescents according to caregiver BP status (Table S1). Adolescents with caregivers who had elevated and hypertensive range BP had a 32% higher prevalence of hypertensive range BP compared with those whose caregivers had normotensive range BP (PR=1.32, 95%CI 1.07; 1.64, p=0.010). This association remained largely unchanged after adjustment for study site and caregiver age in Model 2 (PR=1.32, 95%CI 1.06; 1.65, p=0.012), indicating that caregiver BP is an independent correlate of adolescent BP risk.

**Table 3.**
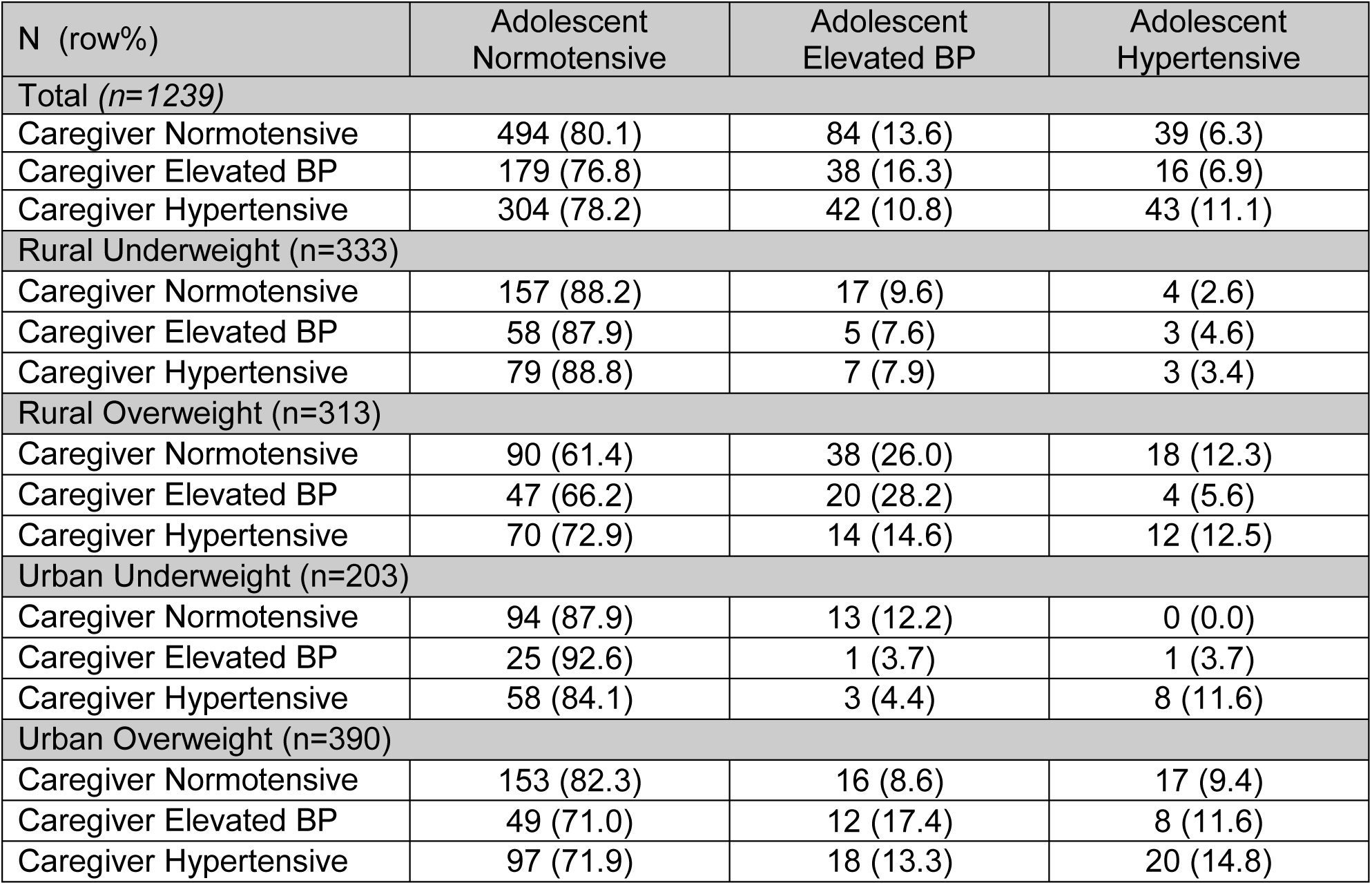
Concordance of blood pressure classification between adolescents and their caregivers.

**Table S1.**
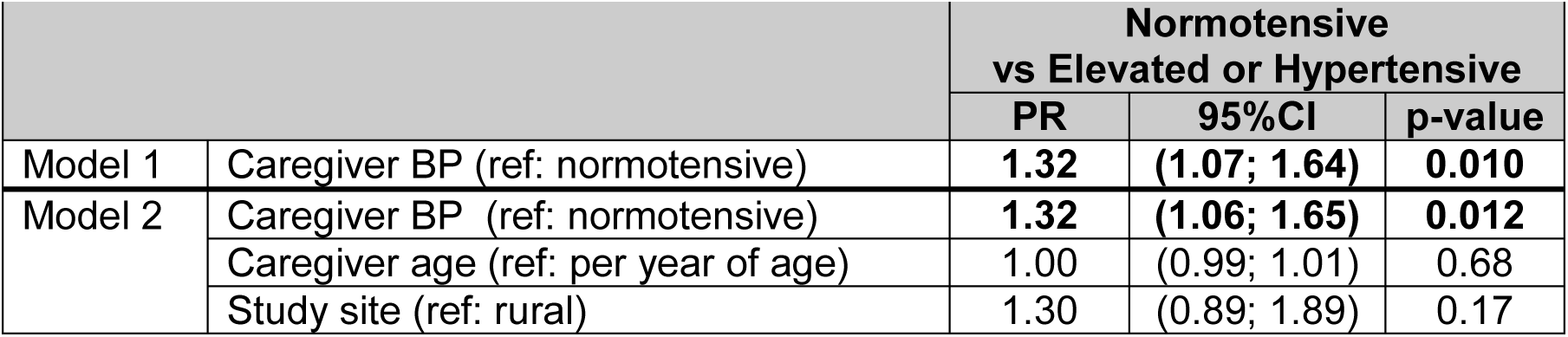
Poisson regression examining the association between caregiver and adolescent blood pressure status.

**Table 4.**
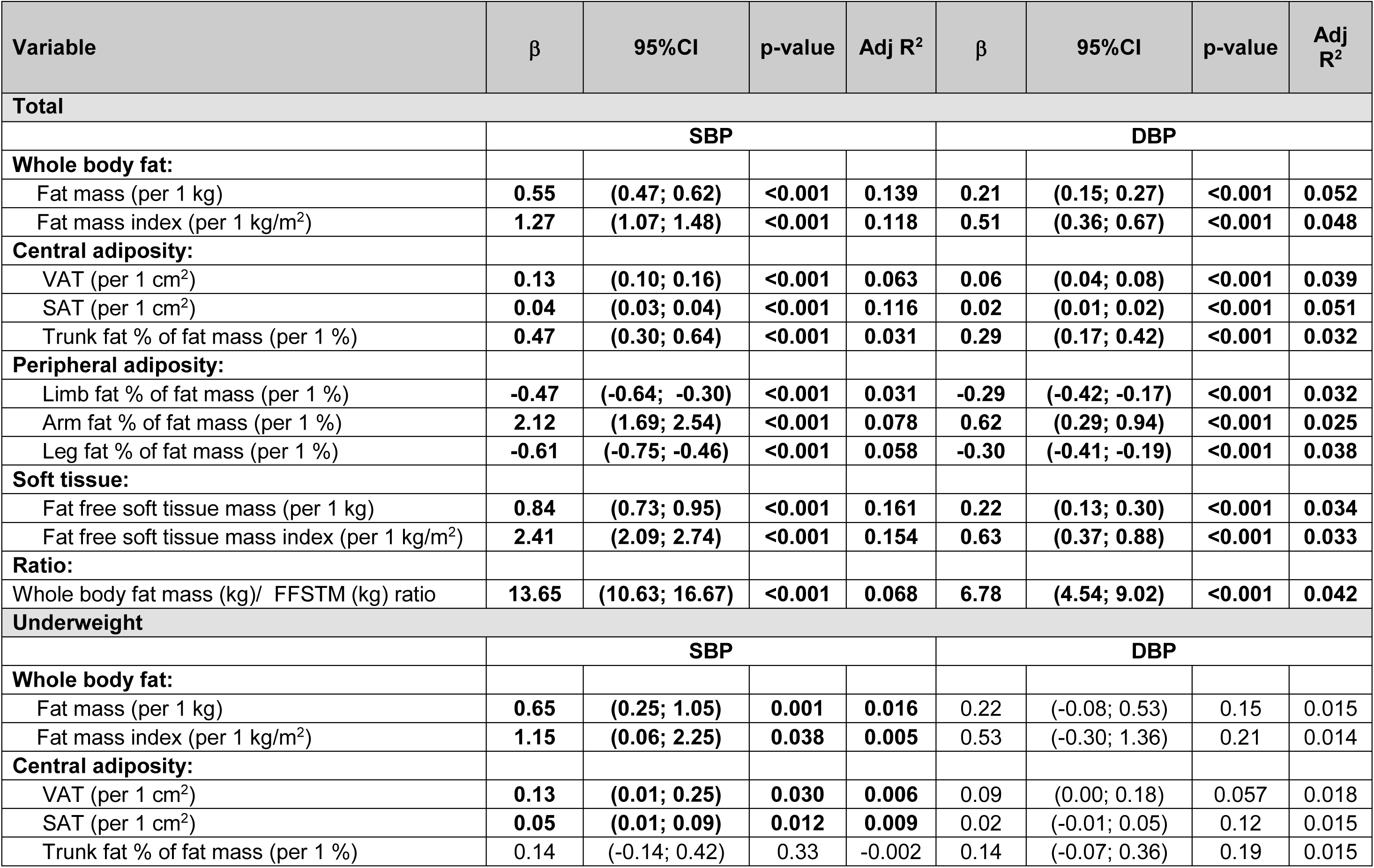
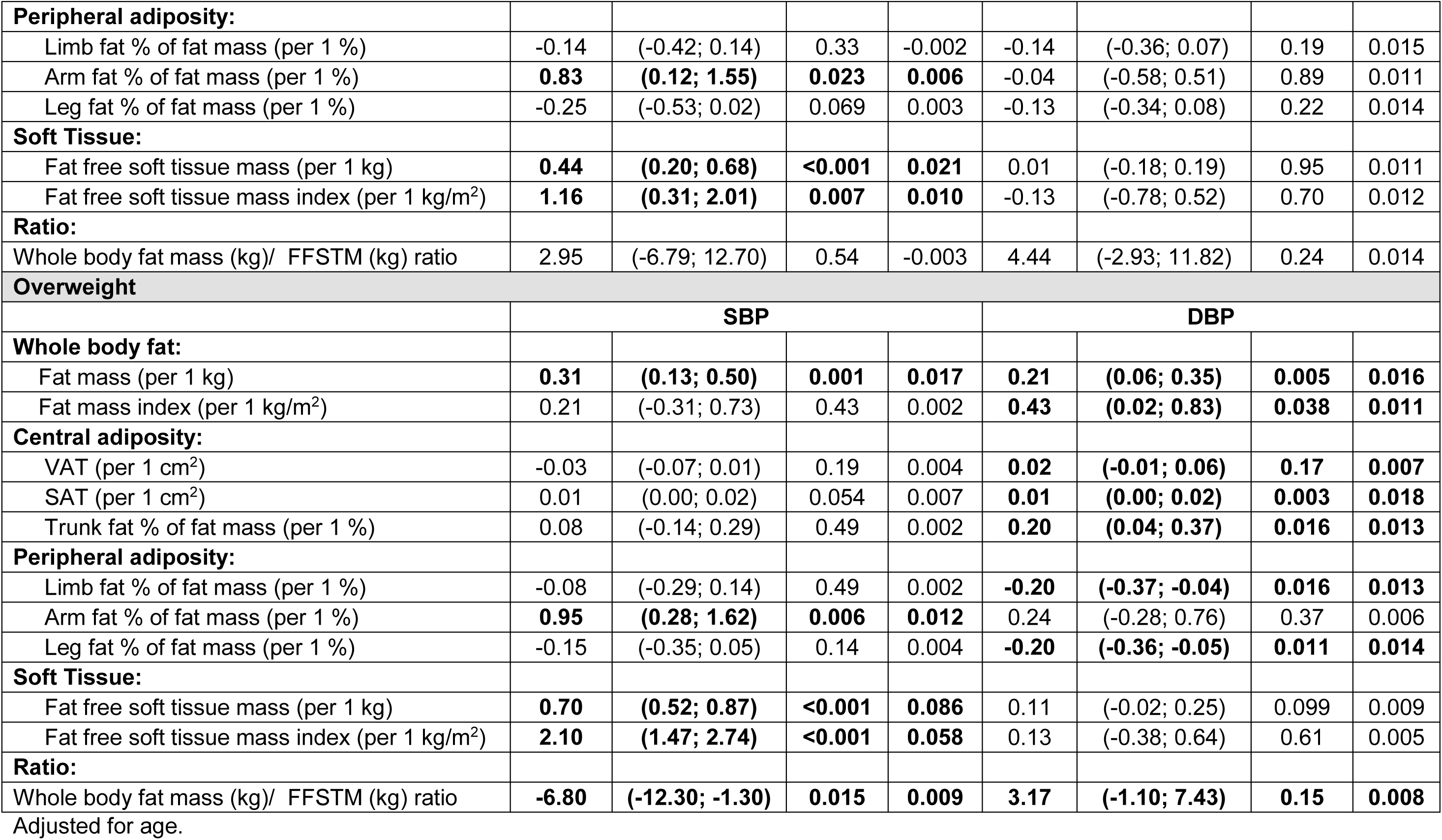
Associations between discrete DXA body composition indices and blood pressure measures (SBP and DBP), adjusted for age.

| Variable | $\beta$ | 95%CI | p-value | Adj R<sup>2</sup> | $\beta$ | 95%CI | p-value | Adj R<sup>2</sup> |
| --- | --- | --- | --- | --- | --- | --- | --- | --- |
| **Total** |  |  |  |  |  |  |  |  |
|  | **SBP** |  |  |  | **DBP** |  |  |  |
| **Whole body fat:** |  |  |  |  |  |  |  |  |
| Fat mass (per 1 kg) | **0.55** | **(0.47; 0.62)** | **<0.001** | **0.139** | **0.21** | **(0.15; 0.27)** | **<0.001** | **0.052** |
| Fat mass index (per 1 kg/m<sup>2</sup>) | **1.27** | **(1.07; 1.48)** | **<0.001** | **0.118** | **0.51** | **(0.36; 0.67)** | **<0.001** | **0.048** |
| **Central adiposity:** |  |  |  |  |  |  |  |  |
| VAT (per 1 cm<sup>2</sup>) | **0.13** | **(0.10; 0.16)** | **<0.001** | **0.063** | **0.06** | **(0.04; 0.08)** | **<0.001** | **0.039** |
| SAT (per 1 cm<sup>2</sup>) | **0.04** | **(0.03; 0.04)** | **<0.001** | **0.116** | **0.02** | **(0.01; 0.02)** | **<0.001** | **0.051** |
| Trunk fat % of fat mass (per 1 %) | **0.47** | **(0.30; 0.64)** | **<0.001** | **0.031** | **0.29** | **(0.17; 0.42)** | **<0.001** | **0.032** |
| **Peripheral adiposity:** |  |  |  |  |  |  |  |  |
| Limb fat % of fat mass (per 1 %) | **-0.47** | **(-0.64; -0.30)** | **<0.001** | **0.031** | **-0.29** | **(-0.42; -0.17)** | **<0.001** | **0.032** |
| Arm fat % of fat mass (per 1 %) | **2.12** | **(1.69; 2.54)** | **<0.001** | **0.078** | **0.62** | **(0.29; 0.94)** | **<0.001** | **0.025** |
| Leg fat % of fat mass (per 1 %) | **-0.61** | **(-0.75; -0.46)** | **<0.001** | **0.058** | **-0.30** | **(-0.41; -0.19)** | **<0.001** | **0.038** |
| **Soft tissue:** |  |  |  |  |  |  |  |  |
| Fat free soft tissue mass (per 1 kg) | **0.84** | **(0.73; 0.95)** | **<0.001** | **0.161** | **0.22** | **(0.13; 0.30)** | **<0.001** | **0.034** |
| Fat free soft tissue mass index (per 1 kg/m<sup>2</sup>) | **2.41** | **(2.09; 2.74)** | **<0.001** | **0.154** | **0.63** | **(0.37; 0.88)** | **<0.001** | **0.033** |
| **Ratio:** |  |  |  |  |  |  |  |  |
| Whole body fat mass (kg)/ FFSTM (kg) ratio | **13.65** | **(10.63; 16.67)** | **<0.001** | **0.068** | **6.78** | **(4.54; 9.02)** | **<0.001** | **0.042** |
| **Underweight** |  |  |  |  |  |  |  |  |
|  | **SBP** |  |  |  | **DBP** |  |  |  |
| **Whole body fat:** |  |  |  |  |  |  |  |  |
| Fat mass (per 1 kg) | **0.65** | **(0.25; 1.05)** | **0.001** | **0.016** | **0.22** | **(-0.08; 0.53)** | **0.15** | **0.015** |
| Fat mass index (per 1 kg/m<sup>2</sup>) | **1.15** | **(0.06; 2.25)** | **0.038** | **0.005** | **0.53** | **(-0.30; 1.36)** | **0.21** | **0.014** |
| **Central adiposity:** |  |  |  |  |  |  |  |  |
| VAT (per 1 cm<sup>2</sup>) | **0.13** | **(0.01; 0.25)** | **0.030** | **0.006** | **0.09** | **(0.00; 0.18)** | **0.057** | **0.018** |
| SAT (per 1 cm<sup>2</sup>) | **0.05** | **(0.01; 0.09)** | **0.012** | **0.009** | **0.02** | **(-0.01; 0.05)** | **0.12** | **0.015** |
| Trunk fat % of fat mass (per 1 %) | **0.14** | **(-0.14; 0.42)** | **0.33** | **-0.002** | **0.14** | **(-0.07; 0.36)** | **0.19** | **0.015** |
| **Peripheral adiposity:** |  |  |  |  |  |  |  |  |
| Limb fat % of fat mass (per 1 %) | -0.14 | (-0.42; 0.14) | 0.33 | -0.002 | -0.14 | (-0.36; 0.07) | 0.19 | 0.015 |
| Arm fat % of fat mass (per 1 %) | **0.83** | **(0.12; 1.55)** | **0.023** | **0.006** | -0.04 | (-0.58; 0.51) | 0.89 | 0.011 |
| Leg fat % of fat mass (per 1 %) | -0.25 | (-0.53; 0.02) | 0.069 | 0.003 | -0.13 | (-0.34; 0.08) | 0.22 | 0.014 |
| **Soft Tissue:** |  |  |  |  |  |  |  |  |
| Fat free soft tissue mass (per 1 kg) | **0.44** | **(0.20; 0.68)** | **<0.001** | **0.021** | 0.01 | (-0.18; 0.19) | 0.95 | 0.011 |
| Fat free soft tissue mass index (per 1 kg/m<sup>2</sup>) | **1.16** | **(0.31; 2.01)** | **0.007** | **0.010** | -0.13 | (-0.78; 0.52) | 0.70 | 0.012 |
| **Ratio:** |  |  |  |  |  |  |  |  |
| Whole body fat mass (kg)/ FFSTM (kg) ratio | 2.95 | (-6.79; 12.70) | 0.54 | -0.003 | 4.44 | (-2.93; 11.82) | 0.24 | 0.014 |
| **Overweight** |  |  |  |  |  |  |  |  |
|  | **SBP** |  |  |  | **DBP** |  |  |  |
| **Whole body fat:** |  |  |  |  |  |  |  |  |
| Fat mass (per 1 kg) | **0.31** | **(0.13; 0.50)** | **0.001** | **0.017** | **0.21** | **(0.06; 0.35)** | **0.005** | **0.016** |
| Fat mass index (per 1 kg/m<sup>2</sup>) | 0.21 | (-0.31; 0.73) | 0.43 | 0.002 | **0.43** | **(0.02; 0.83)** | **0.038** | **0.011** |
| **Central adiposity:** |  |  |  |  |  |  |  |  |
| VAT (per 1 cm<sup>2</sup>) | -0.03 | (-0.07; 0.01) | 0.19 | 0.004 | **0.02** | **(-0.01; 0.06)** | **0.17** | **0.007** |
| SAT (per 1 cm<sup>2</sup>) | 0.01 | (0.00; 0.02) | 0.054 | 0.007 | **0.01** | **(0.00; 0.02)** | **0.003** | **0.018** |
| Trunk fat % of fat mass (per 1 %) | 0.08 | (-0.14; 0.29) | 0.49 | 0.002 | **0.20** | **(0.04; 0.37)** | **0.016** | **0.013** |
| **Peripheral adiposity:** |  |  |  |  |  |  |  |  |
| Limb fat % of fat mass (per 1 %) | -0.08 | (-0.29; 0.14) | 0.49 | 0.002 | **-0.20** | **(-0.37; -0.04)** | **0.016** | **0.013** |
| Arm fat % of fat mass (per 1 %) | **0.95** | **(0.28; 1.62)** | **0.006** | **0.012** | 0.24 | (-0.28; 0.76) | 0.37 | 0.006 |
| Leg fat % of fat mass (per 1 %) | -0.15 | (-0.35; 0.05) | 0.14 | 0.004 | **-0.20** | **(-0.36; -0.05)** | **0.011** | **0.014** |
| **Soft Tissue:** |  |  |  |  |  |  |  |  |
| Fat free soft tissue mass (per 1 kg) | **0.70** | **(0.52; 0.87)** | **<0.001** | **0.086** | 0.11 | (-0.02; 0.25) | 0.099 | 0.009 |
| Fat free soft tissue mass index (per 1 kg/m<sup>2</sup>) | **2.10** | **(1.47; 2.74)** | **<0.001** | **0.058** | 0.13 | (-0.38; 0.64) | 0.61 | 0.005 |
| **Ratio:** |  |  |  |  |  |  |  |  |
| Whole body fat mass (kg)/ FFSTM (kg) ratio | **-6.80** | **(-12.30; -1.30)** | **0.015** | **0.009** | **3.17** | **(-1.10; 7.43)** | **0.15** | **0.008** |

Adjusted for age.

**Table 5.** Multinomial logistic regression between discrete body composition indices and blood pressure classification.

|  | Elevated BP<br>vs Normotensive |  |  | Hypertensive vs Normotensive |  |  |
| --- | --- | --- | --- | --- | --- | --- |
|  | RRR | 95%CI | p-value | RRR | 95%CI | p-value |
| **Total** |  |  |  |  |  |  |
| **Whole Body Fat:** |  |  |  |  |  |  |
| Fat mass (per 1 kg) | **1.06** | **(1.03; 1.08)** | **<0.001** | **1.10** | **(1.07; 1.13)** | **<0.001** |
| Fat mass index (per 1 kg/m<sup>2</sup>) | **1.14** | **(1.07; 1.20)** | **<0.001** | **1.26** | **(1.17; 1.36)** | **<0.001** |
| **Central Adiposity:** |  |  |  |  |  |  |
| VAT (per 1 cm<sup>2</sup>) | **1.01** | **(1.00; 1.02)** | **0.032** | **1.03** | **(1.02; 1.04)** | **<0.001** |
| SAT (per 1 cm<sup>2</sup>) | **1.00** | **(1.00; 1.01)** | **<0.001** | **1.01** | **(1.00; 1.01)** | **<0.001** |
| Trunk fat % of fat mass (per 1 %) | **1.03** | **(0.99; 1.08)** | **0.14** | **1.11** | **(1.05; 1.17)** | **<0.001** |
| **Peripheral Adiposity:** |  |  |  |  |  |  |
| Limb fat % of fat mass (per 1 %) | 0.97 | (0.93; 1.01) | 0.14 | **0.90** | **(0.86; 0.95)** | **<0.001** |
| Arm fat % of fat mass (per 1 %) | **1.39** | **(1.23; 1.57)** | **<0.001** | **1.27** | **(1.10; 1.48)** | **<0.001** |
| Leg fat % of fat mass (per 1 %) | **0.94** | **(0.91; 0.98)** | **0.002** | **0.90** | **(0.86; 0.94)** | **<0.001** |
| **Soft Tissue:** |  |  |  |  |  |  |
| Fat free soft tissue mass (per 1 kg) | **1.10** | **(1.06; 1.13)** | **<0.001** | **1.09** | **(1.05; 1.14)** | **<0.001** |
| Fat free soft tissue mass index (per 1 kg/m<sup>2</sup>) | **1.32** | **(1.20; 1.45)** | **<0.001** | **1.34** | **(1.19; 1.51)** | **<0.001** |
| **Ratio:** |  |  |  |  |  |  |
| Whole body fat mass (kg)/ FFSTM (kg) ratio | **3.49** | **(1.60; 7.60)** | **0.002** | **16.06** | **(6.02; 42.84)** | **<0.001** |
| **Underweight** |  |  |  |  |  |  |
| **Whole Body Fat:** |  |  |  |  |  |  |
| Fat mass (per 1 kg) | 1.05 | (0.92; 1.20) | 0.44 | 1.05 | (0.86; 1.28) | 0.61 |
| Fat mass index (per 1 kg/m<sup>2</sup>) | 1.16 | (0.81; 1.67) | 0.42 | 1.14 | (0.66; 1.98) | 0.64 |
| **Central Adiposity:** |  |  |  |  |  |  |
| VAT (per 1 cm<sup>2</sup>) | 1.00 | (0.96; 1.04) | 0.90 | 1.05 | (0.99; 1.11) | 0.097 |
| SAT (per 1 cm<sup>2</sup>) | 1.00 | (0.99; 1.02) | 0.62 | 1.00 | (0.98; 1.02) | 0.74 |
| Trunk fat % of fat mass (per 1 %) | 0.97 | (0.89; 1.07) | 0.57 | 1.06 | (0.92; 1.23) | 0.38 |
| **Peripheral Adiposity:** |  |  |  |  |  |  |
| Limb fat % of fat mass (per 1 %) | 1.02 | (0.93; 1.13) | 0.57 | 0.94 | (0.81; 1.08) | 0.38 |
| Arm fat % of fat mass (per 1 %) | 1.04 | (0.82; 1.33) | 0.74 | 1.17 | (0.81; 1.70) | 0.40 |
| Leg fat % of fat mass (per 1 %) | 1.02 | (0.93; 1.12) | 0.67 | 0.92 | (0.79; 1.06) | 0.24 |
| **Soft Tissue:** |  |  |  |  |  |  |
| Fat free soft tissue mass (per 1 kg) | 0.98 | (0.90; 1.06) | 0.64 | 1.01 | (0.89; 1.14) | 0.91 |
| Fat free soft tissue mass index (per 1 kg/m<sup>2</sup>) | 0.87 | (0.65; 1.18) | 0.38 | 1.01 | (0.65; 1.58) | 0.96 |
| **Ratio:** |  |  |  |  |  |  |
| Whole body fat mass (kg)/ FFSTM (kg) ratio | 6.97 | (0.30; 159.96) | 0.22 | 2.68 | (0.02; 358.88) | 0.69 |
| **Overweight** |  |  |  |  |  |  |
| **Whole Body Fat:** |  |  |  |  |  |  |
| Fat mass (per 1 kg) | 1.03 | (0.98; 1.07) | 0.23 | **1.08** | **(1.03; 1.14)** | **0.002** |
| Fat mass index (per 1 kg/m<sup>2</sup>) | 1.00 | (0.88; 1.13) | 0.98 | **1.21** | **(1.05; 1.40)** | **0.009** |
| **Central Adiposity:** |  |  |  |  |  |  |
| VAT (per 1 cm<sup>2</sup>) | 0.99 | (0.98; 1.00) | 0.054 | **1.01** | **(1.00; 1.03)** | **0.009** |
| SAT (per 1 cm<sup>2</sup>) | 1.00 | (1.00; 1.00) | 0.68 | **1.01** | **(1.00; 1.01)** | **0.002** |
| Trunk fat % of fat mass (per 1 %) | 1.00 | (0.95; 1.05) | 0.90 | 1.05 | (0.99; 1.11) | 0.12 |
| **Peripheral Adiposity:** |  |  |  |  |  |  |
| Limb fat % of fat mass (per 1 %) | 1.00 | (0.95; 1.06) | 0.90 | 0.95 | (0.90; 1.01) | 0.12 |
| Arm fat % of fat mass (per 1 %) | **1.39** | **(1.17; 1.65)** | **<0.001** | 1.00 | (0.82; 1.22) | 0.98 |
| Leg fat % of fat mass (per 1 %) | 0.97 | (0.93; 1.02) | 0.32 | 0.96 | (0.90; 1.02) | 0.15 |
| **Soft Tissue:** |  |  |  |  |  |  |
| Fat free soft tissue mass (per 1 kg) | **1.10** | **(1.06; 1.16)** | **<0.001** | 1.05 | (1.00; 1.10) | 0.071 |
| Fat free soft tissue mass index (per 1 kg/m<sup>2</sup>) | **1.39** | **(1.18; 1.64)** | **<0.001** | 1.10 | (0.91; 1.34) | 0.33 |
| **Ratio:** |  |  |  |  |  |  |
| Whole body fat mass (kg)/ FFSTM (kg) ratio | 0.29 | (0.08; 1.13) | 0.076 | 3.58 | (0.78; 16.31) | 0.10 |

RRR Relative Risk Ratio

Multivariable linear regression models evaluating the associations of DXA-derived body composition measures with SBP and DBP are presented in **Table 4**. All body composition measures were significantly associated with SBP. Among these, FFSTM explained more of the variance in BP (16.1%) than the other variables. Notably, peripheral fat distribution showed an inverse association with BP. Limb fat as a proportion of total FM was negatively associated with both SBP (β=-0.47, 95% CI:-0.636−-0.301, p<0.001) and DBP (β=-0.29, 95%CI:-0.416−-0.172, p<0.001). This inverse association was driven primarily by leg fat (SBP: β=−0.61, p<0.001, DBP: β=-0.30, p<0.001), whereas arm fat as a proportion of total fat mass was positively associated with both SBP (β=2.12, p<0.001) and DBP (β=0.62, p<0.001). Trunk and limb fat were expressed as complementary proportions of whole-body fat mass and therefore represent a common distributional contrast rather than independent fat depots. A greater proportion of fat located in the trunk, relative to the limbs, was associated with higher SBP and DBP. Conversely, the inverse coefficient for limb fat should be interpreted as reflecting a less central distribution of total fat, rather than as evidence that limb fat is independently protective. The regional findings are therefore consistent with a more central pattern of fat deposition being associated with a less favourable BP profile. Measures of adiposity, including total FM, FMI, and trunk fat percentage, were also associated with SBP, although their contributions to explained variance were generally lower than those of FFSTM indicators. In contrast, body composition variables explain less of the variance in DBP than SBP. Among underweight adolescents, higher FM (β=0.65, p=0.001), FFSTM (β=0.44, p<0.001) and FFSTI (β=1.16, p=0.007) remained positively associated with SBP, and arm fat was also positively associated (β=0.83, p=0.023). Among overweight adolescents, FFSTM (β=0.70, p<0.001) and FFSTI (β=2.10, p<0.001) showed the strong associations with SBP while the FM/FFSTM ratio was inversely associated with SBP (β=-6.80, p=0.015).

Multinomial logistic regression models evaluated the associations between DXA-derived body composition measures and BP classification, adjusted for age (**Table 5**). Across all models, measures of whole body and central adiposity, and FFSTM were strongly and consistently associated with an increased risk of elevated and hypertensive range BP. Measures of whole-body adiposity demonstrated significant associations, with higher FM and FMI associated with increased risk of both elevated (RRR 1.06–1.14) and hypertensive (RRR 1.10–1.26) (all p<0.001) range BP. Central adiposity measures showed similar patterns. Higher FFSTM and FFSTI were associated with increased risk of both elevated and hypertensive range BP. Lower-limb fat showed an inverse association with BP classification. Higher leg fat as a proportion of total fat mass was associated with reduced risk of both elevated (RRR=0.94, 95%CI:0.91–0.98, p=0.002) and hypertensive-range BP (RRR=0.90, 95%CI:0.86–0.94, p<0.001). Arm fat percentage showed the opposite pattern, with higher values associated with increased risk of both elevated (RRR=1.39, 95%CI:1.23–1.57, p<0.001) and hypertensive-range BP (RRR=1.27, 95%CI:1.10– 1.48, p<0.001). When stratified by nutritional status, associations were substantially weaker and mostly non-significant among underweight adolescents, in whom no body composition measure was significantly associated with either elevated or hypertensive-range BP. Among overweight adolescents, adiposity measures remained associated with hypertensive-range BP, including FM (RRR=1.08, 95%CI: 1.03–1.14, p=0.002), FMI (RRR=1.21, 95%CI: 1.05–1.40, p=0.009), VAT (RRR=1.01, 95%CI: 1.00–1.03, p=0.009) and SAT (RRR=1.01, 95%CI: 1.00–1.01, p=0.002). For elevated BP within the overweight group, FFSTM (RRR=1.10, 95% CI: 1.06–1.16, p<0.001), FFSTI (RRR=1.39, 95% CI: 1.18–1.64, p<0.001) and arm fat percentage (RRR=1.39, 95% CI: 1.17–1.65, p<0.001) showed strong associations.

To assess the independent contributions of adiposity and lean mass to BP, a mutually adjusted linear regression model including both FMI and FFSTI, adjusted for age and study site, was fitted (**Table 6**). Both FMI (β=0.74, 95%CI: 0.42–1.06, p<0.001) and FFSTI (β=1.49, 95%CI: 0.97–2.02, p<0.001) were independently and positively associated with SBP. Residing at the urban study site was associated with lower SBP (β=−1.63, 95%CI: −3.17 to −0.08, p=0.039). For DBP, FMI remained independently associated (β=0.50, 95%CI: 0.26–0.75, p<0.001), whereas FFSTI was not (β=0.05, 95%CI: −0.36–0.47, p=0.80). Age was positively associated with DBP (β=0.44, 95%CI: 0.12–0.75, p=0.007). This indicates that both adiposity and lean mass independently contribute to SBP, whereas DBP is driven primarily by adiposity when both are considered simultaneously. We additionally ran the models with FMI and FFSTI standardised and adjusted for age and site, and the findings were similar (results not shown). Both FMI (β = 2.26) and FFSTI (β = 2.69) were associated with systolic blood pressure, while only FMI was associated with diastolic blood pressure (β = 1.54).

**Table 6.** Multivariate linear regression examining the associations between DXA body composition indices and blood pressure measures (SBP and DBP)

| Variable | SBP |  |  | DBP |  |  |
| --- | --- | --- | --- | --- | --- | --- |
| | $\beta$ | 95%CI | p-value | $\beta$ | 95%CI | p-value |
| Fat mass index(per 1 kg/m<sup>2</sup>) | **0.74** | **(0.42; 1.06)** | **<0.001** | **0.50** | **(0.26; 0.75)** | **<0.001** |
| Fat free soft tissue mass index (per 1 kg/m<sup>2</sup>) | **1.49** | **(0.97; 2.02)** | **<0.001** | 0.05 | (-0.36; 0.47) | 0.80 |
| Age | 0.30 | (-0.10; 0.71) | 0.14 | **0.44** | **(0.12; 0.75)** | **0.007** |
| Study site (ref: Rural) | **-1.63** | **(-3.17; -0.08)** | **0.039** | -0.36 | (-1.57; 0.85) | 0.54 |

Sequential multivariable linear regression models were used to evaluate the associations between adolescents’ household, behavioural, caregiver, and body composition factors with SBP and DBP (**Table 7**). In Model 1, higher age was positively associated with SBP (β=1.33, p=0.016), while higher household SES (β=−0.61, p=0.046) and greater household crowding (β=−0.56, p=0.039) were associated with lower SBP. In Model 2, adolescent BMI emerged as a strong predictor of SBP, with overweight adolescents having significantly higher SBP (β=6.75, p<0.001). Age remained positively associated, while behavioural factors were not significantly associated. In Model 3 (caregiver factors alone), neither caregiver BP, caregiver BMI, nor caregiver age were significantly associated with adolescent SBP or DBP. In models combining demographic, household, and behavioural factors (Model 4), BMI remained a strong predictor (β=6.40, p<0.001), and higher household SES remained inversely associated with SBP (β=−0.70, p=0.017). After further adjustment for caregiver factors (Model 5), the association for BMI persisted (β=6.84, p<0.001), while caregiver characteristics were not independently associated with SBP. In the fully adjusted models incorporating DXA-derived body composition (Model 6 and 7 respectively), FMI and FM to FFSTM ratio (FM/FFSTM) were positively associated with SBP (Model 6: β=1.02, p>0.001; Model 7:β=12.05, p=0.008), indicating a strong contribution of body composition to SBP. In this model, household SES remained inversely associated with SBP (β=−0.83, p=0.009). Poor sleep quality was also associated with lower SBP (β=−7.38, p=0.046). For DBP, age was consistently positively associated across all models, from Model 1 (β=1.38, p<0.001) through to the fully adjusted model (Model 6: β=1.45, p=0.001). In Model 2, overweight adolescents had higher DBP compared to underweight adolescents (β=3.06, p=0.010), and this association remained significant in Models 4 and 5 (β≈2.65–2.72, p<0.05). In contrast to SBP, household and behavioural factors were not significantly associated with DBP across models. In the fully adjusted models (Model 6 and 7 respectively), FM/FFSTM was not independently associated with DBP (Model 7:p=0.30). However, poor sleep quality emerged as a significant predictor, associated with lower DBP (Model 6: β=-5.76, p=0.028; Model 7:β=−5.98, p=0.023).

**Table 7.**
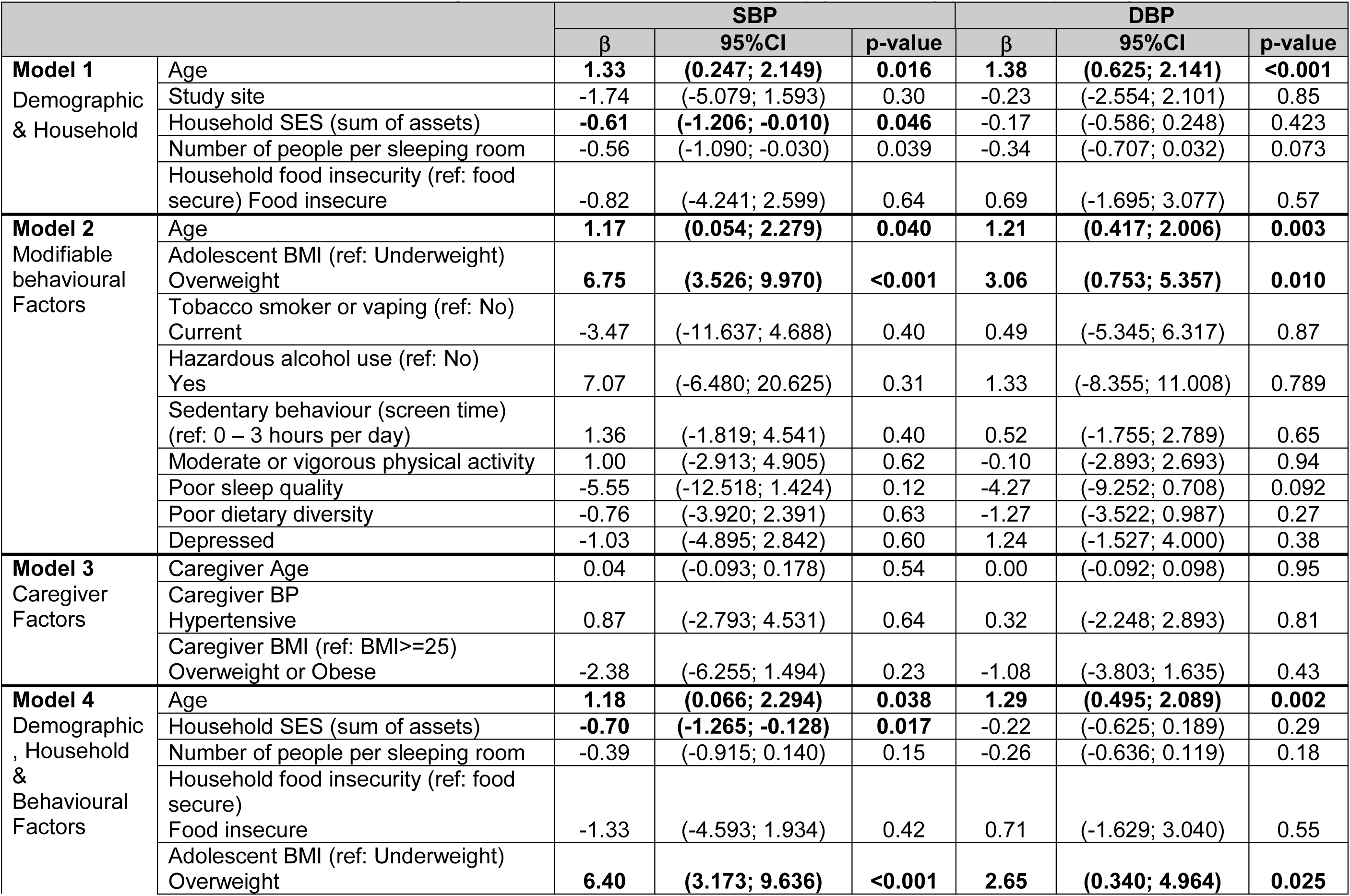
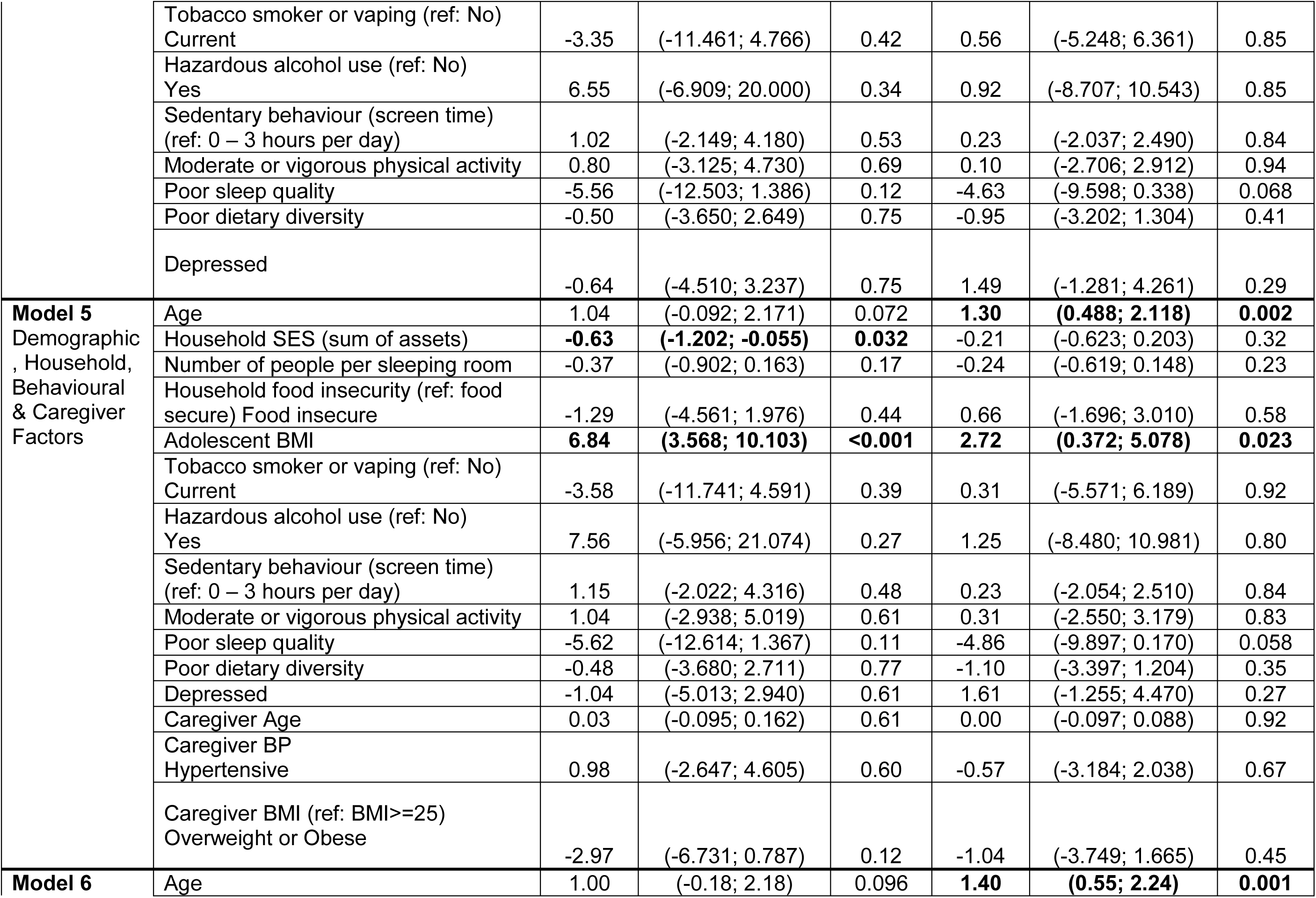
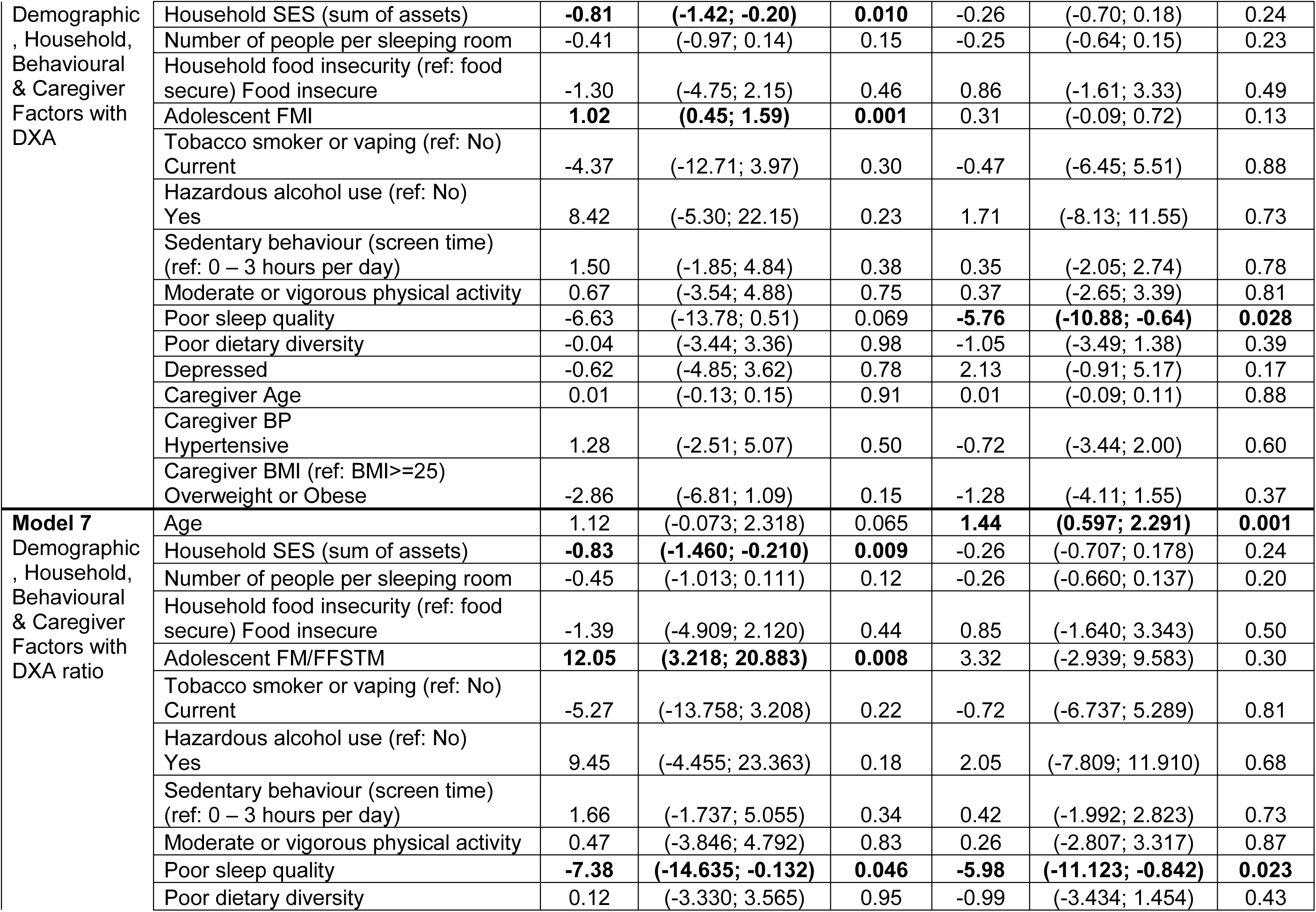
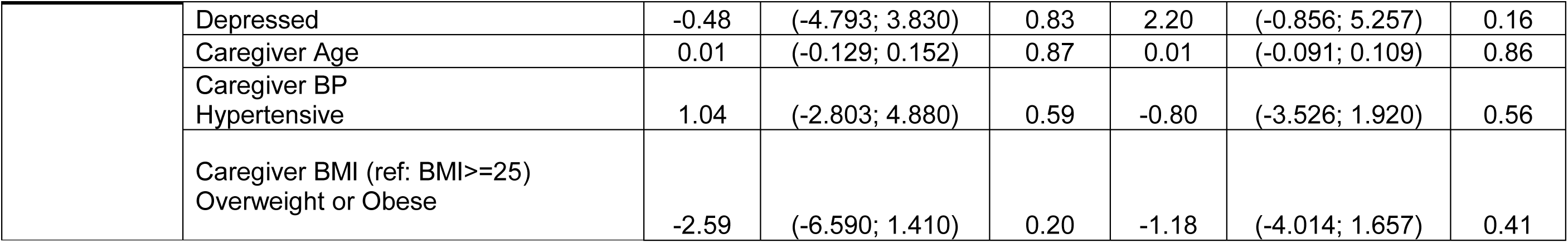
Associations of adolescent and caregiver characteristics measures with (A) SBP and (B) DBP, respectively.

Sequential multinomial logistic regression models evaluated the associations between demographic, household, behavioural, caregiver, and body composition factors with adolescent BP classification (**Table 8**). In Model 1, demographic and household characteristics showed limited associations with BP classification. Residing at the urban study site was associated with a lower likelihood of elevated range BP (RRR=0.66, 95%CI: 0.46–0.96, p=0.030), but not hypertensive range BP. Greater household crowding (number of people per sleeping room) was inversely associated with elevated range BP (RRR≈0.89–0.90, p≤0.010 across models), while no consistent association was observed for hypertensive range BP. Household SES and food insecurity were not associated with BP classification. In Model 2, adolescent BMI emerged as a strong predictor of BP classification. Compared to underweight adolescents, those classified as overweight had more than twice the risk of elevated range BP (RRR=2.52, 95%CI: 1.74–3.63, p<0.001) and approximately four-fold higher risk of hypertensive range BP (RRR=3.99, 95%CI: 2.39–6.66, p<0.001). High sedentary behaviour (>3 hours/day) was associated with an increased likelihood of elevated range BP (RRR≈1.66–1.71, p≤0.010 across models), but not hypertensive range BP.

**Table 8.**
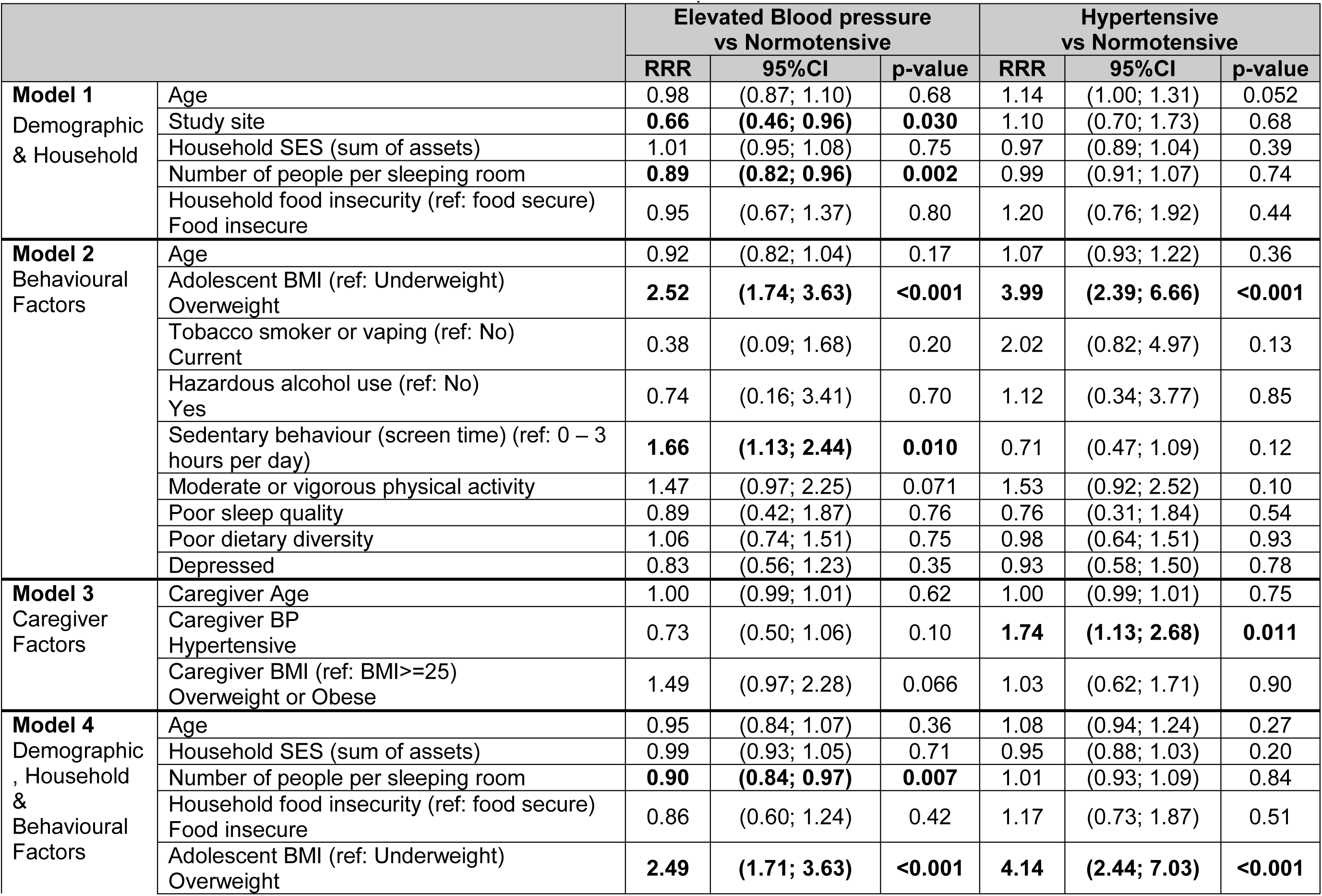
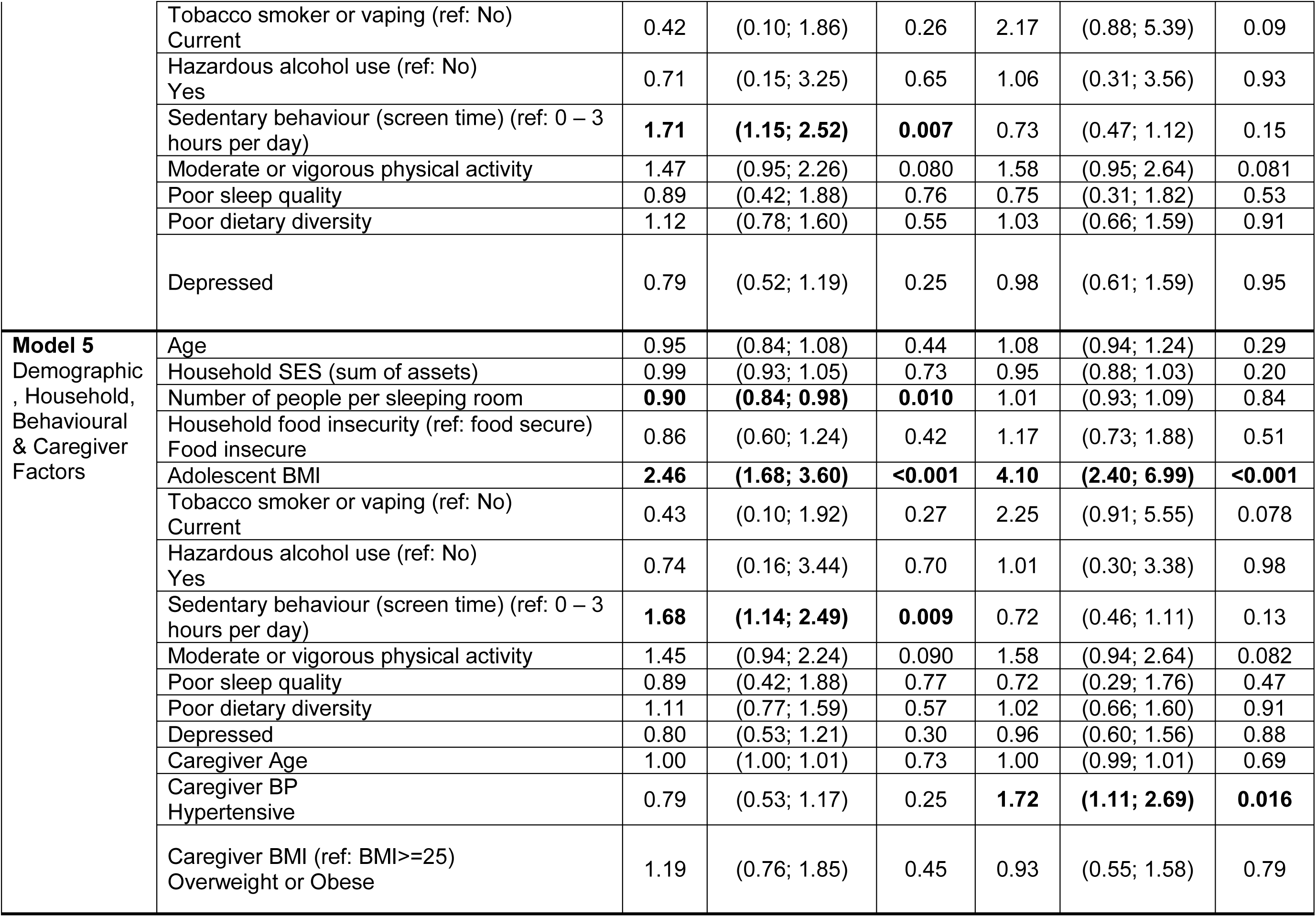
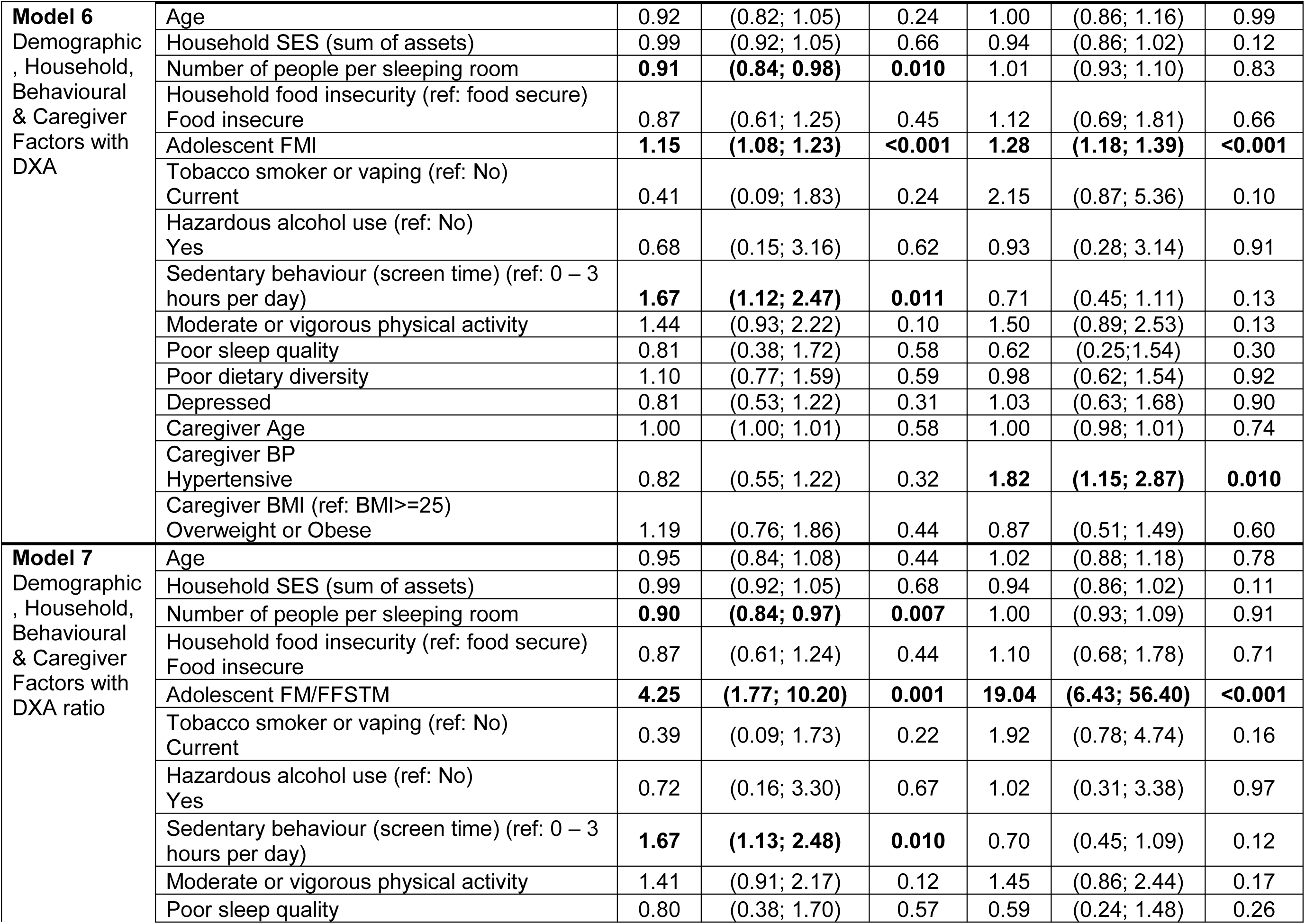
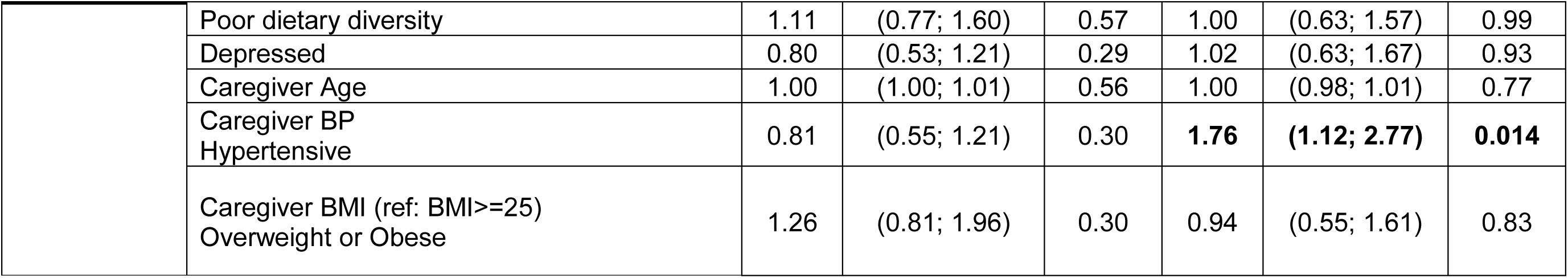
Associations of various characteristics with adolescent blood pressure classification.

Other behavioural factors, including tobacco use, alcohol use, sleep quality, dietary diversity, and depression, were not associated with BP classification. In Model 3, caregiver characteristics were assessed. Caregiver hypertensive range BP was associated with increased risk of adolescent hypertensive range BP (RRR=1.74, 95%CI: 1.13–2.68, p=0.011), and this association persisted in fully adjusted models (Model 5: RRR=1.72, p=0.016; Model 6: RRR=1.76, p=0.014). Caregiver BMI demonstrated a borderline association with elevated range BP (RRR=1.49, p=0.066) but was not associated with hypertensive range BP. Caregiver age was not associated with either outcome. In models incorporating demographic, household, behavioural, and caregiver factors (Models 4 and 5), the associations remained consistent. Overweight adolescents continued to have significantly higher risk of both elevated (RRR≈2.46–2.49, p<0.001) and hypertensive range BP (RRR≈4.10–4.14, p<0.001). Sedentary behaviour remained independently associated with elevated range BP, while household crowding continued to show an inverse association. No additional behavioural or socioeconomic factors emerged as significant predictors. In the fully adjusted models incorporating DXA-derived body composition (Model 6 and 7), Model 6 found higher FMI was independently and strongly associated with a greater risk of both elevated (RRR=1.15, 95%CI: 1.08-1.23, p<0.001) and hypertensive range BP (RRR=1.28, 95%CI: 1.18- 1.39, p<0.001). Indicating that the association between adiposity and higher BP persisted, and was strengthened, when body fat was captured more precisely than by BMI alone. In this model, caregiver hypertensive range BP remained independently associated with adolescent hypertensive range BP (RRR=1.82, 95%CI: 1.15-2.87, p=0.010), sedentary behaviour remained associated with elevated range BP (RRR=1.67, 95%CI: 1.12-2.47, p=0.011), and greater household crowding remained inversely associated with elevated range BP (RRR=0.91, 95%CI: 0.84–0.98, p=0.010). While in Model 7 FM/FFSTM was very strongly associated with BP classification. Higher FM/FFSTM was associated with substantially increased risk of elevated (RRR=4.25, 95%CI:1.77–10.20, p=0.001) and hypertensive range BP (RRR=19.04, 95%CI:6.43– 56.40, p<0.001). Importantly, even after inclusion of body composition, caregiver hypertensive range BP remained independently associated with adolescent hypertensive range BP, and sedentary behaviour remained associated with elevated range BP. The persistence of a statistically significant association across multiple models indicates that this finding is robust and independent of other covariates included in the analyses.

## Discussion

In this large cohort of South African adolescent girls, we show that body composition is a key correlate of BP, with both adiposity and FFSTM independently associated with BP across multiple analytical approaches. Importantly, lower-limb fat demonstrated an inverse association with BP. Among adolescent girls selected according to their BMI status, in rural and urban South Africa, DXA-derived adiposity and FFSTM were associated with higher BP, with consistently stronger and more robust associations observed for SBP than DBP. These findings highlight the value of DXA-derived body composition measures in identifying adolescents at increased cardiometabolic risk beyond BMI alone.

Our findings align with and extend a growing body of evidence demonstrating that cardiovascular regulation in adolescence is influenced by multiple components of body composition acting through distinct physiological pathways. Adiposity, particularly central fat accumulation, contributes to elevated BP through a combination of haemodynamic, endocrine, and inflammatory mechanisms.^29–32^ Excess adipose tissue promotes chronic low-grade inflammation and insulin resistance, which in turn, impairs endothelial function and reduces nitric oxide bioavailability, leading to increased vascular tone.^29–32^ In parallel, adiposity activates the sympathetic nervous system and the renin-angiotensin-aldosterone system, resulting in vasoconstriction, sodium retention, and expansion of intravascular volume.^29–32^ These combined effects increase peripheral resistance and cardiac preload, ultimately elevating BP, particularly SBP. ^29–32^ Consistent with this, longitudinal adolescent studies have shown that higher FM is associated with increased arterial stiffness and early vascular ageing, even before overt hypertension develops.^33^

The positive association between FFSTM and BP warrants careful interpretation. This does not imply that gaining lean mass is detrimental to cardiovascular health. Rather, it reflects the well-established physiological principle that greater metabolically active tissue mass requires increased cardiac output to maintain adequate perfusion.^34^ Larger skeletal muscle mass is associated with greater blood volume, stroke volume, and consequently higher S BP as a normal haemodynamic adaptation to body size.^8^ This is analogous to the observation that taller individuals have higher BP, reflecting physiological scaling rather than pathology. Accordingly, lean mass should be interpreted within the context of overall body composition rather than as an isolated cardiovascular risk factor. A notable feature of our findings was the differential association of the two body composition compartments with SBP and DBP. Adiposity measures, including FM and FMI, were associated with both SBP and DBP, whereas FFSTM was associated predominantly with SBP and showed little independent association with DBP. A larger mass of metabolically active lean tissue increases blood volume, stroke volume, and cardiac output, these changes elevate systolic pressure, whereas diastolic pressure is governed more by peripheral vascular resistance and arterial tone.^8, 35^ Adiposity influences both, contributing to systolic load through increased cardiac output and body size while also raising diastolic pressure via heightened peripheral resistance, sympathetic and renin-angiotensin-aldosterone activation, and adiposity-related vascular dysfunction.^29–32^ The greatest cardiovascular risk is likely to arise when adiposity increases disproportionately relative to FFSTM. When FFSTM increases alongside appropriate fat mass, the haemodynamic adaptation remains within a healthy range. However, when FM accrues disproportionately relative to FFSTM, the combined effects of metabolic dysregulation from adiposity and the physiological demand of existing lean tissue converge to produce a greater net rise in BP. Consistent with this, both FMI and FFSTM were independently and positively associated with BP, and the FM/FFSTM ratio was also strongly associated with BP classification (RRR=4.25 for elevated and RRR=19.04 for hypertensive-range BP). These ratio estimates should not, however, be read as evidence that the FM/FFSTM ratio is the single strongest predictor of BP. The relative risk ratios for the different body composition indices are expressed per their native units (for example, per 1.0 unit of the FM/FFSTM ratio versus per 1 kg/m² of FMI or FFSTI), which span very different ranges, so their magnitudes are not directly comparable. On this like-for-like basis the FM/FFSTM ratio is therefore may not be demonstrably stronger than its individual components; rather, it is a biologically informative composite that captures the balance between adipose and lean tissue and confers cardiovascular risk broadly comparable to that of the separate compartments. Adolescents with a higher proportion of fat relative to lean tissue, that is, those who carry excess adiposity without commensurate muscle mass, represent a particularly high-risk phenotype.^8^ Thus, the FM/FFSTM ratio may be more informative for cardiovascular risk stratification than either FM or FFSTM assessed in isolation, and reliance on BMI alone may obscure meaningful variation in cardiovascular risk. These observations are especially relevant in settings undergoing rapid nutritional transition. By design, this study recruited adolescents at in two different nutritional groups, providing an opportunity to compare the relationships between body composition and BP across distinct phenotypes. As normal-weight adolescents were not included, the design cannot establish whether either extreme carries excess risk relative to the population as a whole; rather, it characterises how body composition relates to BP within and between these two groups. Within this selected sample, overweight adolescents had the highest BP, associated with greater adiposity and higher FM/FFSTM ratios, whereas among underweight adolescents BP was associated more with FFSTM. These cross-sectional patterns are consistent with the view that BP relates to the balance between lean and fat mass rather than to a single component, although longitudinal data with a normal-weight comparison group are needed to confirm whether either extreme confers increased cardiovascular risk.^3, 14^

Sedentary behaviour emerged as an independent and modifiable contributor to the relative risk of elevated BP. Prolonged sedentary time is associated with reduced skeletal muscle activity, impaired glucose uptake, and decreased endothelial responsiveness, all of which can contribute to increased vascular resistance and BP.^36^ Evidence in adolescents indicates that high sedentary behaviour are associated with poorer vascular function and higher BP.^37, 38^ Importantly, the association observed in our study persisted after accounting for body composition, suggesting that sedentary behaviour may influence BP through mechanisms beyond adiposity alone. These findings suggest that behavioural interventions may influence BP through both direct vascular effects and indirect effects on body composition via lifestyle. Reducing sedentary time during adolescence may therefore represent a practical target for early CVD prevention, particularly in populations experiencing the double burden of malnutrition.

The association between caregiver and adolescent hypertensive range BP highlights the importance of intergenerational risk. This likely reflects shared genetic susceptibility (where caregivers are biologically related) as well as common environmental exposures, including diet, physical activity, and household stressors.^39, 40^ Early-life exposure to these factors may “programme” cardiovascular regulation, increasing susceptibility to elevated BP during adolescence, consistent with the developmental origins of health and disease (DOHaD) framework.^41^ These findings support the need for household-level or family-based prevention strategies, rather than approaches focused solely on individuals.

This study must be interpreted by its strengths and limitations. Key strengths include the large sample size, inclusion of both urban and rural populations, and the use of gold-standard body composition measures. The integration of behavioural, household, and caregiver data enabled a comprehensive analysis of multilevel correlates. Limitations include the cross-sectional design, which precludes causal inference, and potential residual confounding, including from unmeasured factors such as dietary sodium intake. BP was assessed at a single visit, which may overestimate hypertensive range BP due to measurement variability; confirmatory repeat measurements and longitudinal follow-up are needed to determine whether these clinic BP patterns predict persistent hypertension and later cardiovascular risk. Behavioural measures were self-reported and may be subject to misclassification. The analyses examining DXA-derived body composition measures across multiple BP outcomes were exploratory in nature, and no correction for multiple comparisons was applied. While the majority of associations were statistically robust at conventional thresholds, the possibility of type I error cannot be excluded, and findings should be interpreted in the context of the overall pattern of results rather than reliance on any single comparison. Confirmatory studies with pre-specified hypotheses are warranted. Interaction tests were exploratory, and likely underpowered within the smaller strata.

At the individual level, the results highlight the need for early-life interventions targeting healthy body composition, specifically, reducing excess and central adiposity while supporting lean mass accrual and linear growth through adequate nutrition and physical activity, rather than implying that FFSTM gain alone will lower BP. Given that sedentary behaviour was independently associated with elevated BP, strategies to reduce prolonged screen time and promote regular moderate-to-vigorous physical activity during adolescence represent modifiable targets. At the household level, the independent association between caregiver and adolescent BP underscores the importance of household-centred approaches that simultaneously address cardiometabolic risk across generations. Interventions delivered through community health workers that engage both adolescents and their caregivers may be particularly effective in resource-limited settings where clinic-based follow-up is inconsistent. Screening caregivers for hypertension during adolescent health contacts could also identify high-risk household units for targeted prevention. Finally, the use of DXA-based measures strengthens the evidence that body composition is a key factor linking BMI status to early cardiovascular risk.^42^

In conclusion, body composition was associated with BP among underweight and overweight adolescent girls, particularly for SBP. Greater adiposity and FFSTM were positively associated with SBP, while regional percentage analyses were consistent with higher BP when fat was distributed more centrally. The FM/FFSTM ratio provided complementary information about relative body composition but was not demonstrably superior to FMI or FFSTI. Longitudinal studies incorporating repeat BP measurements, a normal-weight comparison group and appropriately modelled regional body composition are needed to assess the persistence and clinical relevance of these associations.

## Data Availability

The data are available from the corresponding author upon reasonable request.

## Acknowledgements

The SAMRC/Wits Rural Public Health and Health Transitions Research Unit and Agincourt Health and socio-Demographic Surveillance System, a node of the South African Population Research Infrastructure Network (SAPRIN), is supported by the Department of Science, Technology and Innovation, the University of the Witwatersrand, and the Medical Research Council, South Africa, and previously the Wellcome Trust, UK (grants 058893/Z/99/A; 069683/Z/02/Z; 085477/Z/08/Z; 085477/B/08/Z). The SAMRC/Wits Developmental Pathways for Health Research Unit by the South African Medical Research Council and the University of the Witwatersrand, Johannesburg, South Africa. SJS and KKO are supported by the UKRI Medical Research Council (MC_UU_00006/2) and the NIHR Cambridge Biomedical Research Centre (NIHR203312).

## Author Contributions

All authors were involved in the conception and planning of the study. SHC carried out the data analyses, generated figures and tables and interpreted the data .SHC did the literature search and the writing of the paper. AC, LKM, NC, SC, EM, KKO, SMT, KK and SHC read and contributed to the final version. All authors provided edits and approved the final version.

## Data availability

The data are available from the corresponding author upon reasonable request.

## References

1. Khoury M, Urbina EM. Hypertension in adolescents: diagnosis, treatment, and implications. The Lancet Child & Adolescent Health. 2021;5(5):357–66.

2. Norris SA, Frongillo EA, Black MM, et al. Nutrition in adolescent growth and development. The lancet. 2022;399(10320):172–84.

3. Crouch S, Mathatha D, Micklesfield LK, et al. Describing the triple burden of malnutrition in adolescents in rural and urban South Africa. South Afr J Clin Nutr. 2025:1–7.

4. Pomeroy E, Macintosh A, Wells JC, et al. Relationship between body mass, lean mass, fat mass, and limb bone cross-sectional geometry: Implications for estimating body mass and physique from the skeleton. American Journal of Physical Anthropology. 2018;166(1):56–69.

5. Diaz-Canestro C, Pentz B, Sehgal A, et al. Lean body mass and the cardiovascular system constitute a female-specific relationship. Sci Transl Med. 2022;14(667):eabo2641.

6. Han TS, Al-Gindan YY, Govan L, et al. Associations of body fat and skeletal muscle with hypertension. The Journal of Clinical Hypertension. 2019;21(2):230–8.

7. Wells Jonathan CK. Using Body Composition Assessment to Evaluate the Double Burden of Malnutrition. Ann Nutr Metab. 2019;75(2):103–8.

8. Yu S, Zhao S, Tang J, et al. Fat-free mass may play a dominant role in the association between systolic blood pressure and body composition in children and adolescents. British Journal of Nutrition. 2024;131(4):622–9.

9. Jones LL, Griffiths PL, Norris SA, et al. Is puberty starting earlier in urban South Africa? American Journal of Human Biology: The Official Journal of the Human Biology Association. 2009;21(3):395–7.

10. Lundeen E, Norris S, Adair L, et al. Sex differences in obesity incidence: 20-year prospective cohort in S outh A frica. Pediatr Obes. 2016;11(1):75–80.

11. Micklesfield LK, Pedro TM, Kahn K, et al. Physical activity and sedentary behavior among adolescents in rural South Africa: levels, patterns and correlates. BMC Public Health. 2014;14(1):40.

12. Matshipi M, Monyeki K. Longitudinal relationship between skinfolds ratios and blood pressure from childhood to late adolescence among Ellisras rural children: Ellisras Longitudinal Study. African Journal for Physical Activity and Health Sciences (AJPHES). 2024;30(2):254–67.

13. Monyeki K, Kemper H, Makgae P. The association of fat patterning with blood pressure in rural South African children: the Ellisras Longitudinal Growth and Health Study. Int J Epidemiol. 2006;35(1):114–20.

14. Burrows R, Correa-Burrows P, Reyes M, et al. Low muscle mass is associated with cardiometabolic risk regardless of nutritional status in adolescents: A cross-sectional study in a Chilean birth cohort. Pediatr Diabetes. 2017;18(8):895–902.

15. Kahn K, Collinson MA, Gómez-Olivé FX, et al. Profile: Agincourt health and socio-demographic surveillance system. Int J Epidemiol. 2012;41(4):988–1001.

16. Richter L, Norris S, Pettifor J, et al. Cohort profile: Mandela’s children: the 1990 birth to twenty study in South Africa. Int J Epidemiol. 2007;36(3):504–11.

17. Norris SA, Draper CE, Prioreschi A, et al. Building knowledge, optimising physical and mental health and setting up healthier life trajectories in South African women (Bukhali): a preconception randomised control trial part of the Healthy Life Trajectories Initiative (HeLTI). BMJ open. 2022;12(4):e059914.

18. Castell GS, Rodrigo CP, de la Cruz JN, et al. Household food insecurity access scale (HFIAS). Nutr Hosp. 2015;31(3):272–8.

19. World Health Organisation. The Alcohol Use Disorders Identification Test (AUDIT) [Available from: https://www.who.int/publications/i/item/WHO-MSD-MSB-01.6a.

20. Buysse DJ, Reynolds III CF, Monk TH, et al. The Pittsburgh Sleep Quality Index: a new instrument for psychiatric practice and research. Psychiatry Res. 1989;28(2):193–213.

21. Kroenke K, Spitzer RL, Williams JB. The PHQ-9: validity of a brief depression severity measure. J Gen Intern Med. 2001;16(9):606–13.

22. Food and Agriculture Organization (FAO). Individual Dietary Diversity. 2014.

23. World Health Organisation. Global School-based Student Health Survey 2021 [Available from: https://cdn.who.int/media/docs/default-source/ncds/ncd-surveillance/gshs/gshs_core_expanded_questions_2021.pdf?sfvrsn=8878627_3.

24. Cole TJ, Lobstein T. Extended international (IOTF) body mass index cut-offs for thinness, overweight and obesity. Pediatr Obes. 2012;7(4):284–94.

25. Flynn JT, Kaelber DC, Baker-Smith CM, et al. Clinical practice guideline for screening and management of high blood pressure in children and adolescents. Pediatrics. 2017;140(3).

26. Unger T, Borghi C, Charchar F, et al. 2020 International Society of Hypertension global hypertension practice guidelines. Hypertension. 2020;75(6):1334–57.

27. Harris PA, Taylor R, Minor BL, et al. The REDCap consortium: building an international community of software platform partners. Journal of biomedical informatics. 2019;95:103208.

28. Harris PA, Taylor R, Thielke R, et al. Research electronic data capture (REDCap)—a metadata-driven methodology and workflow process for providing translational research informatics support. Journal of biomedical informatics. 2009;42(2):377–81.

29. Cassis LA, Police SB, Yiannikouris F, et al. Local adipose tissue renin-angiotensin system. Curr Hypertens Rep. 2008;10(2):93–8.

30. Schütten MT, Houben AJ, de Leeuw PW, et al. The link between adipose tissue renin-angiotensin-aldosterone system signaling and obesity-associated hypertension. Physiology. 2017;32(3):197–209.

31. Blueher M. An overview of obesity-related complications: The epidemiological evidence linking body weight and other markers of obesity to adverse health outcomes. Diabetes, Obesity and Metabolism. 2025;27:3–19.

32. Karakasis P, Stachteas P, Iliakis P, et al. Inflammation and Resolution in Obesity-Related Cardiovascular Disease. Int J Mol Sci. 2026;27(1):535.

33. Dangardt F, Charakida M, Georgiopoulos G, et al. Association between fat mass through adolescence and arterial stiffness: a population-based study from The Avon Longitudinal Study of Parents and Children. The Lancet Child & Adolescent Health. 2019;3(7):474–81.

34. Grijalva-Eternod CS, Lawlor DA, Wells JC. Testing a capacity-load model for hypertension: disentangling early and late growth effects on childhood blood pressure in a prospective birth cohort. PLoS One. 2013;8(2):e56078.

35. DeMers D, Wachs D. Physiology, mean arterial pressure. StatPearls [internet]: StatPearls Publishing; 2023.

36. Kerr NR, Booth FW. Contributions of physical inactivity and sedentary behavior to metabolic and endocrine diseases. Trends Endocrinol Metab. 2022;33(12):817–27.

37. Böhm B, Kirchhuebel H, Elmenhorst J, et al. Sedentary behavior in childhood, lower arterial compliance and decreased endothelial function-cross sectional data from a German school cohort. Frontiers in Pediatrics. 2022;9:787550.

38. de Moraes ACF, Carvalho HB, Rey-López JP, et al. Independent and combined effects of physical activity and sedentary behavior on blood pressure in adolescents: gender differences in two cross-sectional studies. PLoS One. 2013;8(5):e62006.

39. Wójcik M, Alvarez-Pitti J, Kozioł-Kozakowska A, et al. Psychosocial and environmental risk factors of obesity and hypertension in children and adolescents—a literature overview. Frontiers in cardiovascular medicine. 2023;10:1268364.

40. Ware LJ, Maposa I, Kolkenbeck-Ruh A, et al. Are cardiovascular health measures heritable across three generations of families in Soweto, South Africa? A cross-sectional analysis using the random family method. BMJ open. 2022;12(9):e059910.

41. Tain Y-L, Hsu C-N. Developmental and early life origins of hypertension: preventive aspects of melatonin. Antioxidants. 2022;11(5):924.

42. Tanasescu M-D, Rosu A-M, Minca A, et al. Beyond BMI: Rethinking Obesity Metrics and Cardiovascular Risk in the Era of Precision Medicine. Diagnostics. 2025;15(23):3025.

